# Enhancing Mental Health Decision-Making with Artificial Intelligence/Machine Learning: A Prescriptive Analytics Approach for Customised Outcomes

**DOI:** 10.1101/2025.06.16.25329741

**Authors:** Mark Payne, Fareed Ud Din, Kabir Sattarshetty, Cassandra Sundaraja, Anwaar Ul-Haq, Theresa Scott, Niusha Shafiabady

## Abstract

Depression is a complex and widespread mental health condition affecting over 280 million people globally, yet access to timely diagnosis and personalised treatment remains limited. This study explores the application of artificial intelligence and machine learning (AI/ML) to develop a prescriptive analytics framework for supporting mental health decision-making. Using a dataset of 2,556 anonymized datapoints, the study evaluates multiple machine learning classifiers to identify depression risk, benchmarked by accuracy, F1 score, and AUC metrics. Logistic Regression emerged as the most balanced and interpretable model, achieving an accuracy of 97.3% and an AUC of 99.9%, with the lowest false-negative rate—crucial in a clinical support context. The study then introduces an ensemble modeling framework based on stratified Logistic Regression models, enabling consensus-based predictions and enhanced reliability. Beyond predictive analytics, the framework facilitates prescriptive insights by identifying minimal, targeted lifestyle changes likely to shift individuals from high to low depression risk. Feature importance was established using statistical and effect size measures, guiding personalized intervention suggestions with practical interpretability. The study highlights relationships among key features such as academic pressure, work hours, job satisfaction, and financial stress, revealing compound intervention opportunities. Despite limitations related to dataset provenance and generalizability, the framework demonstrates promising utility in non-clinical mental health contexts and forms a tractable foundation for future deployment in professional settings. These findings underscore the potential of AI/ML to augment mental health care by delivering scalable, explainable, and data-driven decision support tools aimed at improving patient outcomes through personalized, prescriptive strategies.

## Graphical Abstract

## Introduction

Depression is a heterogeneous and multifactorial mental health disorder [1] effecting an estimated 5% of the adult population, or 280 million people, worldwide [2] [3] – this far exceeds the one million mental health practitioners available to combat the illness globally [4].

In recent years, the advent of Artificial Intelligence (AI) and Machine Learning (ML) has revolutionized various fields. The integration of AI/ML into mental health care holds immense potential for improving diagnosis, treatment and overall patient outcomes. By leveraging data, these technologies can uncover patterns and insights that previously laid latent, establishing ways to deliver more personalized and effective mental health care, in ways that are more tractable, scalable, available and effective.

Despite these advancements, a significant research gap remains in understanding the full potential and application of AI/ML in mental health, where applied data science can help to better understand depression by analysing patterns to identify key factors, predict occurrences and prescribe solutions.

To do this we need to account for uncertainty from two main sources [5] – variation within the dataset and variation within the model. Within the dataset, researchers reported challenges obtaining mental health datasets with sufficiently large sample sizes [6], [7],[8], [9], [10] using a variety of approaches to obtain salient data ranging from interviews and questionnaires [11] to controlled studies [12] bolstered by lower barrier sources such as ChatGPT [13] to generate simulated datasets, and text mining of social media posts (Twitter, Reddit, and Facebook) [14],[15], to capture requisite sample sizes, however with the latter classed as “self-reporting” [16] - found to reduce general performance due to “inherent uncertainty…methods fail to capture” [17], and while some canonical mental health data sources/standards do exist (STAR*D, PRISMA, and ARPNET for example) [18] their scope is specifically targeted, therefore not generalisable, while datasets in general were often inherently stratified [13] attributed to influences in responder participation [19] including cost, time pressure, social stigma, gender tendency (females more likely to visit their doctor), and other motivation factors - with strong opportunity for technology to help overcome these barriers.

Compounding this, studies show use of automated recommendations can have significant influence on practitioner decision making [20], making it critical to address variation in the model - ensuring that models are as reliable and explainable as possible, where various factors can impact model accuracy and reliability, ranging from experiment design [20] to recruitment approach [21], to the very nature of classification - where high raw accuracy can give way to low classification precision and specificity [22], important given such a high stakes area. This makes model explainability and simplicity critical [23],[24] to ensure confidence in output results, complicated with lack of agreed/standard definitions of what ‘good’ looks like [25] with performance checks often limited to internal validation alone [26], using predictive correlation over causality [27] lacking statistical significance [28], [29], meaning model coverage is incomplete, therefore likely to cause accuracy drop from testing to leave out and ground truth [30] comparison.

Even with the dataset and model baselined, it raises the question of utility, where meta-studies appear to show the majority of machine learning in mental health research has so far employed predictive analytics, the detection of illness, where very few studies went further into prescriptive analytics, interventions to reduce likelihood of the illness, and when they did their focus was in highly specific remediations, such as quantity of sleep, nutrition or exercise plans[31].

The significance of this study lies in its potential to support mental health care by providing a more robust, transparent, accessible and generalisable framework for prescriptive Analytics based AI/ML applications. By addressing the current practice gaps between data acquisition and modelling, this study aims to enhance the reliability and availability of machine learning based prescriptive mental health models, ultimately leading to better participant outcomes. Additionally, the study will offer broad dynamic treatment approaches and real-world effective decision support systems, helping to overcome aforementioned barriers such as practitioner capacity constraints, cost, patient stigma, and case motivation [32].

Ultimately, this study seeks to bridge the research gap in AI/ML mental health applications by providing comprehensive insights into model performance and decision support tractability. The findings will contribute to the development of more reliable and accessible mental health care solutions, ultimately improving the quality of life for individuals affected by mental health conditions.

## Methods

The target dataset was collected during an anonymous survey of respondents in January–June 2023 drawn from diverse backgrounds and professions across various cities within India, aimed at identifying factors contributing to depression risk among adults aged 18–60. The dataset is publicly available and accessible online from the following source **[dataset][DS]**. Participants provided information on demographic and lifestyle factors including age, gender, city, education, job/study satisfaction, work/study hours, and family history, without requiring clinical mental health assessments.

The target variable, *Depression*, indicates whether an individual is at risk of depression (*Yes* or *No*), based on their responses. The dataset is designed to explore correlations between everyday factors and mental health risks, making it valuable for predictive modelling and research in non-clinical mental health contexts, and as a precursor to use of such models in a clinical context.

Depression is a critical mental health issue therefore data provenance is equally critical. It’s important to call out that in obtaining this dataset, no reference was found to suggest practitioner involvement in data acquisition or assessment. On this basis, the dataset should be considered indicative only, where the cases contained are not representative of expert assessment/practitioner judgement but rather a placeholder for exploring the potential opportunity, with ability to replace and build an authoritative dataset from real world professional input in practice.

While the dataset isn’t authoritative, it is useful to investigate ability to apply a broad case feature set, determine predictive ability related to mental health diagnosis, and ability to capture and leverage latent relationships within the data, for improved decision making and support. Cognisant of this, it’s insightful to perform Exploratory Data Analysis to better understand the dataset before use.

### Exploratory Data Analysis

Exploratory Data Analysis (EDA) is a key stage in the data science life cycle. EDA is the use of quantitative and visual techniques to analyse and inspect the target dataset to better understand the data under observation – to discover patterns, identify anomalies and ultimately gain a better understanding of the structure, distribution, and quality of the data, while assisting to identify incomplete or incorrect datapoints that could negatively impact machine learning model quality, to formulate treatments required.

The target dataset is made up 2,556 data samples containing 19 features - 8 numerical and 11 categorical. The histograms depicted in **Figure 1** below provide a high level view of the numerical features, allowing appreciation of normality and independence of values contained that, with an apparent absence of bell-shaped curve across the board, indicates a strong lack of normality.

**Figure 1:** Distribution of numerical features within the target dataset.

The boxplots in **Figure 2** below provide further insights for Depression representation across numeric features, where a strong inverse relationship appears to exist with *Age* and *Depression*, the ‘No’ group having an IQR of 43 compared to the ‘Yes’ group IQR of 25 and maximum/outliers of 51. This is contrary to empirical evidence where Indian based depression studies tend to show depression significantly increases in later life [[**D**]], especially beyond 60 years of age, likely indicating this dataset covers a specific subset or strata of the Indian population, or that curation has potentially been applied.

**Figure 2:** Box plots of categorical features within the target dataset.

Meanwhile a strong relationship appears to exist for both *Academic Pressure* and *Work Pressure* with higher tendencies of *Depression*, while *CGPA* appears to show no impact on depression at all. Both the *Study Satisfaction* and *Job Satisfaction* boxplots indicate an inverse likelihood of depression, with *Job Satisfaction* slightly less so, while increased *Work/Study Hours* appears to moderately increase likelihood of depression. Finally increased *Financial Stress* appears to increase depression rates.

Evaluating the spread of depression across categorical parameters, the combined graph in **Figure 3** below appears to show *Gender* represents a marginal increase between male and female cases at 18.1% Male vs 17.5% Female. Again, this appears contrary to empirical evidence [33] expecting significantly higher female depression rates compared to male [**B**], suggesting this dataset is non normal and/or somewhat curated - potentially to address reported gender disparity [34].

**Figure 3:** Depression distribution by categorical feature.

Meanwhile *Students* appear to show a significantly higher rate of depression compared to working *Professionals*, interesting given empirical studies (with the younger average age of students) again tend to show the inverse **[D]**. *Sleep duration* appears to show a mild influence on the rate of depression, while *Dietary habits* also appear to influence depression where improved diet shows a reduced rate of depression. The existence of *Suicidal Thoughts* appears to increase the likelihood of depression, while *Family History of Mental Illness* (17.1% with vs 18.6% without) appears to show almost no bearing on depression.

The *City* plot in **Figure 4** (and **Appendix A** for more detail) below appears to show a fair degree of consistency between depression rates within Indian cities however *Hyderabad* stands out with the highest representation of depression (32.9%), followed by Lucknow (22.7), and Sringar (23.5). This compares to the bottom three cities of *Varanasi* (12.0), *Delhi* (12.2), and *Jaipur* (12.5).

**Figure 4:** Depression distribution by City, Degree, and Profession (see Appendix A for detail.)

The Degrees plot in **Figure 4** below appears to show the *Class12* category is a standout with 45.8% of respondents associated with depression. Given the over representation, *Class 12* (the final year of school in India) would attract further attention, while again the young average age (20.4 Years) and prevalence of depression in *Class 12* holders appears contrary to empirical expectations **[D]**. Mindful the majority (62%) of Class 12 Holders are recorded as *Working Professional* indicating they are in the workforce and not degree qualified– therefore in a potentially lower socioeconomic status, compared to other *Working Professionals* that hold a professional degree, potentially coinciding with the higher *Depression* prevalence **[D]**.

Reviewing the relationship between *Professions* for depression, the top three most prevalent appear to be *Graphic Designer* (23.1%), the next a tie between *Judge*, *Data Scientist* and *HR Manager* (14.3%), compared to the least prevalent being *Pharmacist* (1.3%), *Entrepreneur* (1.6%) and *Chemist* (1.7%)

These distributions appear interesting because Indian based depression surveys increasingly report the likelihood of depression increases with age rather than decreasing. Furthermore, the target dataset appears limited to professionals, or soon-to-be professionals in a degree course. Collectively, the divergence of parameters such as *Age* and *Gender* from empirical distributions, and the non-normal nature of the distribution, indicates this dataset is likely a subset of the population, or potentially curated.

As identified in initial data sourcing, the dataset lacks provenance to suggest authoritative practitioner involvement or clinical controls. While this dataset is no less useful for evaluating automated diagnosis and prescriptive analytics, the lack of normal distribution and stratification to higher socioeconomic population would otherwise impact ability to generalise result from this study, if the source was captured in clinical conditions.

Finally, the histogram in **Figure 5** below guides parameter importance with respect to depression, highlighting the most influential parameters measured by statistical significance (influence) and effect size (magnitude), measuring numerical feature importance (p-value) using the *Mann-Whitney U Test* and the effect size guided by *Cohen’s d* comparing the standard deviations between features. For categorical variables, the *Chi-Square* Test is used to assess the association between categorical variables and depression, and *Cramér’s V* to calculate the effect magnitude in each relationship. Used together both sets of methods provide a balanced guide to both significance and practical relevance.

**Figure 5:** Effect size and importance by feature.

Evaluating these metrics, *Age*, with an effect size of 1.450, is the most influential feature and significantly associated with depression (P<0.05) followed by *Financial Stress* (effect size 0.439.) While *Academic Pressure* (1.084) and *Work Pressure* (0.681) show moderate to high effect sizes, they aren’t statistically significant, and *Profession* (0.154), *City* (0.111), *CGPA* (0.089), *Family History of Mental Illness* (0.018) and *Gender* (0.007) aren’t statistically significant and show negligible effect sizes.

### Feature Engineering

With EDA completed and with better awareness of the target dataset, we apply a data pre-processing pipeline to prepare the dataset for machine learning. Data engineering treatments applied here include imputing missing data with static values, which is important as many ML algorithms do not know how to handle missing data, scaling of numerical features, where proportionate parameter weighting improves machine learning accuracy, and one-hot encoding of categorical features, to ensure tractable machine learning across subsets. The Depression target variable is also label-encoded.

### Classification Model Selection

We evaluate a range of supervised ML algorithms on our target dataset to determine the best performer. In general, classification algorithms can be represented by a decision function that minimises a particular cost [**A0**], with specific implementations as follows:

- **Logistic Regression** is a classification algorithm that identifies linear relationships between data features and the log-odds of outcomes by converting inputs to probabilities between 0 and 1 through a logistic function, while employing maximum likelihood estimation to determine optimal coefficients that create the most effective separating hyperplane. [**AL1**]
- **CatBoost** is a specialized gradient boosting algorithm optimized for categorical feature handling. It transforms categorical variables into numerical representations during the training process. This ensemble method builds sequential decision trees where each new tree learns from the errors of the previous iterations. CatBoost’s Ordered Boosting technique helps prevent overfitting, resulting in robust and reliable performance. [**AL2**]
- **Support Vector Machine (SVM)** is a supervised learning classifier that identifies the optimal hyperplane the widest possible margin between classes in feature space, utilizing the closest points (support vectors) from each class. Through kernel functions like radial basis or polynomial, SVMs effectively manage non-linear relationships and high-dimensional data. [**AL3**]
- **Neural Network** is an architecture inspired by the biology of the brain. It features multiple layers of connected nodes (neurons) with weighted connections that are adjusted during training. Non-linear activation functions enable complex pattern recognition. The network learns through backpropagation, computing error gradients across weighted connections and updating them via optimization methods like gradient descent. This versatile approach excels at discovering nuanced data patterns. [**AL4**]
- **Gradient Boosting** is an ensemble technique that combines multiple decision trees into a robust predictor. It progressively enhances accuracy by minimizing prediction errors through loss functions such log loss for classification. While capable of handling sophisticated data patterns and supporting custom loss functions, it requires careful regularization to avoid overfitting. [**AL5**]
- **Random Forest** is an ensemble learning approach that creates multiple decision trees, each trained on different data samples and feature subsets. By developing diverse trees affected by different error patterns, random forests minimize the variance inherent in individual trees. For classification tasks, the model aggregates tree predictions through majority voting, producing more reliable and accurate results that generalize well to new data. [**AL6**]
- **Naive Bayes** is a probability-based classifier leveraging Bayes’ Theorem to determine class likelihood given observed features. It’s "naive" in its assumption that features are conditionally independent within each class, which streamlines calculations by computing class posterior probabilities and selecting the most probable outcome. This straightforward approach makes it particularly effective for large, high-dimensional datasets. [**AL7**]

### Training and Testing

Each model is then trained on the target dataset of 2,556 data samples (respondents), with 10 randomly sampled *Depression* records withheld for use in hold back testing, splitting the remaining data 80/20 for use in model training and testing, against the following metrics:

- **Accuracy:** The withheld accuracy indicates how often the model correctly classifies unseen examples.
- **F1 score:** Measures the mean of the precision (proportion of correct positive predictions) vs recall (the proportion of actual positive instances correctly identified.)
- **AUC score:** AUC (Area Under Curve) score evaluates the model’s ability to discern positive and negative classes.

The histogram in **Figure 6** below depicts the relative performance of each algorithm across accuracy, F1 score, and AUC.

**Figure 6:** Comparison of Model performance by Accuracy, F1 Score and AUC score.

Most models perform quite well, with scores generally above 0.8 across all metrics. *Logistic Regression*, despite being one of the simplest models, shows strong performance comparable to more complex models. *Naïve Bayes* performs notably worse than other models, with scores around 0.3-0.5. *CatBoost, XGBoost*, and the *Neural Network* show very consistent performance across all three metrics. The AUC Score (green bars) tends to be slightly higher while Precision and F1 scores for most models.

The performance of each algorithm is tabled and analysed in detail below, including a corresponding confusion matrix for each algorithm, where the green quadrants tally the correctly labelled *Depression* status, while the red quadrants tally the incorrectly labelled cases, where the false negative quadrant (bottom left) forms a key metric in health domains, representing cases where positive cases are misclassified as negative – in our case missed depression cases. This can have serious consequences, therefore orientation to a conservative classification is preferred.

With the best Accuracy [97.27%], F1 Score [92.13%] and AUC [99.86] *Logistic Regression* displayed the most balanced accuracy, precision, and recall, while showing minimal variance in cross-validation and, with high AUC and F1 scores, handled class imbalance effectively.

The more sophisticated models (*CatBoost, SVM, Neural Network*) show better balance between sensitivity and specificity. However, the low number of false negatives in the top performing models is crucial. There’s a reasonable trade-off with false positives (cases incorrectly labelled as having *Depression*) which is generally preferable to the alternative - missing cases in mental health screening. The high AUC scores suggest these models could be valuable tools for initial depression screening.

Looking at the confusion matrix, *Logistic Regression* handled classification the best, with the lowest number of false negative classifications (one compared to the nearest *Catboost’s* with 13) important as a miss-diagnosed *Depression* case is critical in a real-world care scenario. And while *Logistic Regression*’s false negative labels are a little high with 13, you would rather a conservative approach, than the alternative.

The next best performing algorithm, *Catboost*, provided identical accuracy, slightly lower F1 Score and AUC, with expected ability to handle non-linearity and complex data patterns, especially with categorical variables, enabled solid handling of class imbalance. *SVM* displayed high accuracy and AUC but slightly lower F1 scores, showing a good balance between precision and recall though less effective for minority classes detection. The *Neural Network* displayed similar performance to *SVM* with slightly better precision for minority class detection. *Gradient Boosting* achieved good overall accuracy however it struggled with minority class recall, potentially due to class imbalance. *Random Forest* performed similarly to Gradient Boosting, with solid accuracy, but also impacted by minority class recall. *Naive Bayes* drastically underperforms on all metrics, its simplistic model lacked ability to handle non-linear relationships and interdependencies among parameters.

### Hold Back Testing

As identified in previous studies, internal testing doesn’t describe a model’s full predictive power. For this reason and for the benefit of completeness the models are also independently trained and evaluated against 80 randomly withheld cases/respondents (40 with Depression, 40 without.)

As shown in [**Table 2**] below, this guides the ground truth predictive accuracy, where the confusion matrix reinforces that *Logistic Regression*’s hold-back accuracy holds up extremely well. The results are encouraging with high precision and recall, while the F1-score is excellent, demonstrating that the model captures the variability in the data well.

**Table 1:** Model Training Results.

**Table 2:** Ground Truth Hold Out Test Results.

On this basis *Logistic Regression* appears to be the best candidate for deployment, due to strong performance, reliability, simplicity and inherent explainability, with *CatBoost* the fallback contender, if non-linearity or interpretability was a concern. Again, it’s encouraging that *Logistic Regression* has the lowest quantity of false negative classifications, making for a more conservative, lower risk classifier.

### Model Framework

Next, we build a framework to ensemble a set of three *Logistic Regression* models for use in *Depression* prediction. This facilitates representation of one or more practitioner trained models, and with practitioner judgement encoded, allows for a distributed, consensus-based approach to diagnosis.

While we initially ensemble three models, the quantity could be expanded to any number required in a multiple practitioner arrangement. This not only allows visibility of practitioner judgment, otherwise latent in the data, but provides balance across multiple practitioner assessment and consensus, increasing reliability in decision support. Furthermore, whether used in multiple or standalone arrangements, an ensemble approach provides a number of statistical benefits to model quality including:

- Balancing out individual biases (reducing potential for overfitting and making the model less susceptible to noise in the training data) and better enables generalisation of new, unseen data.
- Uncorrelated errors get corrected by consensus, reducing mistakes, providing better stability and more consistent results.
- Less sensitive to outliers and noise, therefore more reliable in production environments.

To do so we apply the data preprocessing pipeline used previously in model evaluation, to three stratified data subsets, ensuring a balanced distribution of depression cases in our multi-practitioner arrangement. Model building and optimisation is undertaken utilising hyperparameter tuning via *RandomizedSearchCV*, and consensus is mediated across the three models using a hard voting mechanism, to aggregate ensemble predictions, where each model outputs a binary vote based on its own trained weighting/probability of depression, with the majority vote determining the final classification.

### Single Feature Interventions

With the ensemble assembled, we are in position to apply prescriptive analytics capabilities to target interventions. Using the feature significance and magnitude that we quantified previously, we train heuristics (rules of thumb) to seek out the ‘lowest barrier’ interventions indicated by the smallest lifestyle change(s) quantified by the magnitude difference required to change the predicted *Depression* outcome from ‘Yes’ to ‘No.’ This is done by fixing to a specific client record, the “As-Is” health assessment (current parameter values) and incrementing through potential “To-Be” parameter values, initially restricting to one discrete change per iteration (though expanding to multiple parameter combinations later for flexibility) enabling visibility of the updated *Depression* propensity, and appreciation of the otherwise latent relationships encoded within model features, to make prescriptive decision support as simple and practical as possible for real world intervention and assistance of client outcomes.

## Results and Discussion

### Outcomes

To evaluate performance the model is first calibrated on classification accuracy, ensuring feature values result in the same *Depression* ‘Yes’ classifications as previously recorded. Once confirmed, we perform prescriptive analytics using the 10 held back *Depression* records, observing ability to reduce/remove depression likelihood via inspecting the resulting performance visuals and logs, captured for practitioner interpretability and insight.

The visuals in **Figure 7** below show the results from automated feature increments to single features, aggregated per case/respondent, showing the impact to *Depression* probability for each parameter value change – old value vs new. The results are encouraging, with multiple visuals showing new “to-be” parameter values close to their original “as-is” values, indicating a small change required to reduce depression to below threshold, further encouraging because all cases appear to transition to a ‘No’ *Depression* prediction outcome.

**Figure 7:** Comparison of single feature interventions (old vs new values) by case.

The utility here is that the visualization shows where remediation efforts are most concentrated - of most help in resource allocation and intervention planning. It also highlights the cases that may require more comprehensive support. Observations include for example that *Work/study Hours* and *Academic Pressure* frequently succeed in intervention, *Job Satisfaction* and *Work Pressure* often present together, suggesting a complementary nature, and *Financial Stress*, while often present, appears to require relative lower magnitude of change to succeed in influencing *Depression* outcomes. Meanwhile, *Academic Pressure* and *Study Satisfaction* tend to present together, and work-related factors (*Hours, Pressure, Satisfaction*) tend to present together, while *Sleep Duration* improvements often appear to coincide with reduced *academic/work pressure*. Cases with high *Academic Pressure* often show corresponding high *Work/Study hours*.

### Relationships

With encouraging results, the question broadens from our initial basis - whether the models show significant and meaningful results, to the question of whether the results are tractable. Rather than rely on single feature changes to influence *Depression* prediction, we can evaluate the potential of taking a combined therapeutic approach – using one, two or more interventions to reduce the probability of *Depression* classification from ‘Yes’ to ‘No’.

It would assist to gain better awareness of the relationships within the features, the graph in **Figure 8** below provides some insight into the size and nature (positive or negative) of relationships between features, describing how each feature interacts with the others, to infer constraints and valid permutations/combinations for automated interventions, and for awareness in general to support practitioner decision making.

**Figure 8:** Statistical relationships between features.

We first confirm the relationships depicted below calibrate with the parameter importance observations made earlier, where the relationship graph appears to show a strong positive correlation between *Academic Pressure* and *Depression*, suggesting academic stress is a significant risk factor, and likewise *Financial Stress* and *Work/Study Hours* match with previous metrics. *Job Satisfaction* shows a negative correlation indicating higher *Job Satisfaction* may help protect against depression. This all appears consistent.

With alignment confirmed, we can focus more deeply on relationships within the data, where *Job Satisfaction* appears to connect strongly to *Work Pressure* and *Professional status*, and *Work/Study Hours* connects to both academic and professional elements, potentially showing how deeply interconnected work-life balance factors are. The presence of *Suicidal Thoughts* shows concerning connections to multiple stressors.

The complex interconnections and multiple interacting factors suggest that combined interventions could be effective to address depression, though the inclusion of multiple features adds complexity, taking on elements of a combinatorial search problem, where the potential search space increases with every feature, and by a factor of complexity with categorical features having multiple labels. Just considering raw feature combinations limited to five numeric and five categorical values respectively, the search space would result in time complexity of O(5^9^ x 5^9^) = O(5^18^) which is in the region of computational intractability.

Heuristics such as feature pruning assist to limit the complexity by slackening the search space, where a potential limitation becomes an advantage. While previous results were promising, further guard rails are required to ensure meaningful recommendations. To do so we apply a set of constraints, the first a set of tractable parameters. This has multiple benefits. For example, *Age* has a large and significant influence on depression, however, there’s no ability to influence an individual’s chronological age. It is however important to respect how age interacts with other features that are tractable for our purposes, and by accounting for the influence in the model, enable more effective interventions using parameters the respondent can influence.

By focussing on features a respondent can influence, we reduce the permutations available and therefore the search space is reduced while the statistical relationships remain respected. Features retained are as follows: *Sleep Duration, Dietary Habits, Working/Student, Financial Stress, Work Pressure, Work/Study Hours, Study Satisfaction, Job Satisfaction, Academic Pressure*, where *Work/Study Pressure* and *Job/Study satisfaction* tend appear mutually exclusive.

We then slacken the search space further by limiting to use of 3 feature combinations, while using early stopping to exit once a resolution is identified at the required level: This reduces the time complexity to become much more manageable.

### Combined Feature Evaluation

With feature constraints defined a fresh set of hold-out samples are chosen at random, and we run the framework to evaluate prescriptive ability. With results summarised below in **Figure 9** this visualization guides how frequently each respective feature is used in each case across all remediations, where the scale 0 to 2 represents the number of times a feature is referenced.

**Figure 9:** Frequency of feature use in remediations.

The visuals show encouraging results where each case has at least one successful single feature intervention, with six cases, including *Case 300*, *561*, and *2242*, can be addressed using multiple combined feature resolutions. Multiple features such as *Financial Stress*, *Work/Study Hours*, and *Dietary Habits* show frequent use across multiple cases, very encouraging as this aligns with the same features ranked highest in our earlier feature importance metrics, while *Academic Pressure* and *Study Satisfaction* show consistent usage patterns across cases. This in contrast to *Working Professional* or *Student status* that shows relatively few mentions. Overall, what appears most encouraging is that each case has at least one single or combined successful intervention.

Next the heat map in **Figure 10** shows the magnitude of intervention in various features across each case studied, where the colour scale ranges from 0 (dark blue) to 12 (deep yellow), indicating the cumulative use of change, where most features show moderate changes across cases (lighter blue shades.) Interesting cases include *Case 561* which requires high-touch interventions across *Financial Stress*, *Job Satisfaction*, *Work Pressure* and *Work/Study hours*, while *Case 1666* shows particular focus on *Work/Study Hours*, *Work Pressure* and *Job Satisfaction*. In terms of features, *Work Study Hours* and *Financial Stress* require the highest value change, while *Working Professional/Student status* and *Dietary Habits* are lowest touch. *Study Satisfaction* and *CGPA* appear to be used least.

**Figure 10:** Magnitude of feature use in remediations.

Most encouraging is that the natural relationships between the data appears to be maintained where, for example, where *Academic Pressure* is only seen in absence of *Job Satisfaction*, while *Job Satisfaction* for example is always used in exclusion of student related features.

Insights from these matrices suggests that while certain features (e.g. *Work/Study Hours*) show both significant magnitude of change and frequent use, others might be used more often and require less change, or vice versa. This could be valuable for understanding which intervention aspects are most attempted versus which actually show meaningful results in the *Depression* outcomes.

Meanwhile the results summarised below in **Figure 11** demonstrates the feature value increments that lead to depression prediction remediation from ‘Yes’ to ‘No’, showing the average influence (probability reduction) vs the average change required (the magnitude.) This indicates the relative ‘return on effort’, the likely benefit from an applied intervention, where the lower the average change and higher the influence the better.

**Figure 11:** Feature use and average value difference.

Interesting observations emerge, for example, *Work/Study Hours* is the most used feature with 13 instances, however with the highest change effort (average change 5.44 out of 12), while *Financial Stress* and *Academic Pressure* are frequently used - 12 and 7 times respectively - with slightly higher intervention effort (∼2.2–2.4 average out of 5), indicating they are highly effective and mid-barrier to address, while *Study Satisfaction* is also the lowest effort among the top used features, making it a highly efficient focus area. Features like *Job Satisfaction* and *Work Pressure* are moderately used (4 times each) with slightly higher efforts (2.5 each.) *Dietary Habits* is used 7 times while *CGPA* is used only once (4) but are efficient to address with average value difference around ∼1. Sleep Duration is the least used (3 times), indicating minimal focus or relevance in interventions, while possibly indicating that sleep duration is an outcome of another factor.

### Combined Feature Interventions – Detailed

Finally, the **Figure 12** below shows feature interventions per case, in order of change effort (magnitude of change.) The range of single and combined feature interventions is encouraging, with 5 cases resolved by one feature change, and the remaining cases resolved by combined interventions. Furthermore, the range of combined interventions provides flexibility where multiple smaller changes can be evaluated instead of a large intervention if otherwise restricted to single features.

**Figure 12:** Magnitude of feature change by case and intervention.

**Figure 13:** Depression distribution by City.

**Figure 14:** Depression distribution by Degree.

**Figure 15:** Depression distribution by Profession.

At this point, in a real-world implementation, practitioners would ideally update the dataset with the latest client outcomes, so the most recent ‘as-is’ details and *Depression* classification match for real world calibration.

Even while constrained to a defined set, the breadth of the features modelled for use in combined interventions demonstrates ability to apply broad horizon mental health insights and interventions, providing a common and transparent view on data, predictions and treatments, while the model explainability and visuals allow multi-disciplinary teams to collaborate more easily, using approaches that are readily externally testable, with feature sets that can be expanded and improved incrementally, providing required transparency to build maturity in and, ultimately, for potential use in clinical trials. Furthermore, this approach has potential to be delivered online in a highly available and scalable way, to help overcome barriers to entry, while supporting practitioner decision support, self-reference, collaborative consensus, and once proven, broader respondent self-service.

## Conclusion

With strong results this study gives confidence that, with a combination of targeted design principles and standards driven approach, we can deliver robust, reliable and scalable data driven prescriptive analytics to help tackle mental health conditions like Depression, and support better mental health outcomes. We highlighted the benefits of AI, and the gaps between current research and practical applications of ML in mental health.

We acknowledged issues compounded by use of automated recommendations, targeting models that were both high performing and highly explainable – with model reliability and transparency critical given the significant influence on practitioner decision making, driven by factors such as experiment design, recruitment approach, classification algorithm transparency and simplicity, while going some way to resolve the lack of agreed/standard definitions of what ‘good’ looks like in mental health machine learning, with performance measured beyond internal validation alone, using predictive correlation, causality and statistical significance, to expand ability to generalise.

In demonstrating this we overcame a number of challenges identified in previous studies: dealt with the uncertainty in the dataset and in the model, identified controls required, addressed factors such as dataset sample size and quality, influence of methods used in data sourcing, and implications for data credibility and confidence, addressing data issues that arise using targeted treatments and techniques, highlighting gaps in standards, otherwise limiting ability to generalise.

Looking forward, the lack of normal distribution and apparent stratification/curation of our target dataset would likely impact ability to generalise, while the lack of provenance means that while we obtained encouraging results, for truly authoritative decision support, real world practitioner collaboration and data capture is critical to build out a broadly tractable feature set, encoding judgement of the highest credibility, for real world evaluation and use. This illustrates the need for defined, authoritative and generalised mental health datasets and standards, to enable diversion of effort from capturing and analysing the data, to focus on advancing the standard and depth of the modelling and decision support available. We hope to have gone some way to demonstrate the value of this effort.

While seeking to build such datasets, we advise to assess the impact that cultural factors can have on mental health, potentially influenced by western and non-western cultural basis, for example where our EDA appeared to identify the target dataset diverged from empirical observations, that could materially impact the veracity of outputs generated. This may invite additional consideration around data handling, potentially addressed with approaches such as regional labelling, segmentation, or partitioning.

And with that, to achieve more generalisable model performance, and to accommodate a dataset with more complex/nuanced relationships, it’s worth considering more advanced algorithms such as *neural networks* (as explainability methods improve) or ensembles such as *Random Forest*, *Catboost*, or *Gradient Boosting*. Mindful this would require further preprocessing to address class imbalance, such as oversampling or other cost-sensitive methods.

Finally, it would be worth evaluating search optimisation for the prescriptive analytics/interventions provided. While we used heuristics to good effect, there’s significant potential for industrial optimisation techniques to improve prescriptive recommendations. For example, reinforcement learning has featured heavily in recent breakthroughs in large language model performance, such as R1, and with ability to sustain the transparent interpretability required for decision making, reinforcement learning could be a strong candidate for our purposes.

As guided in the introduction of this research, the significance of this study lies in the potential for machine learning to support mental health care for detection and treatment of conditions such as depression, to provide a step forward for improved mental health outcomes, to support practitioners with robust, transparent, capable and accessible machine learning and prescriptive analytics, ultimately supporting better mental health outcomes.

## Data Availability

The data underlying the results presented in the study are publicly available from the Kaggle dataset ‘Depression Survey Dataset for Analysis’ (URL: https://www.kaggle.com/datasets/sumansharmadataworld/depression-surveydataset-for-analysis).

---

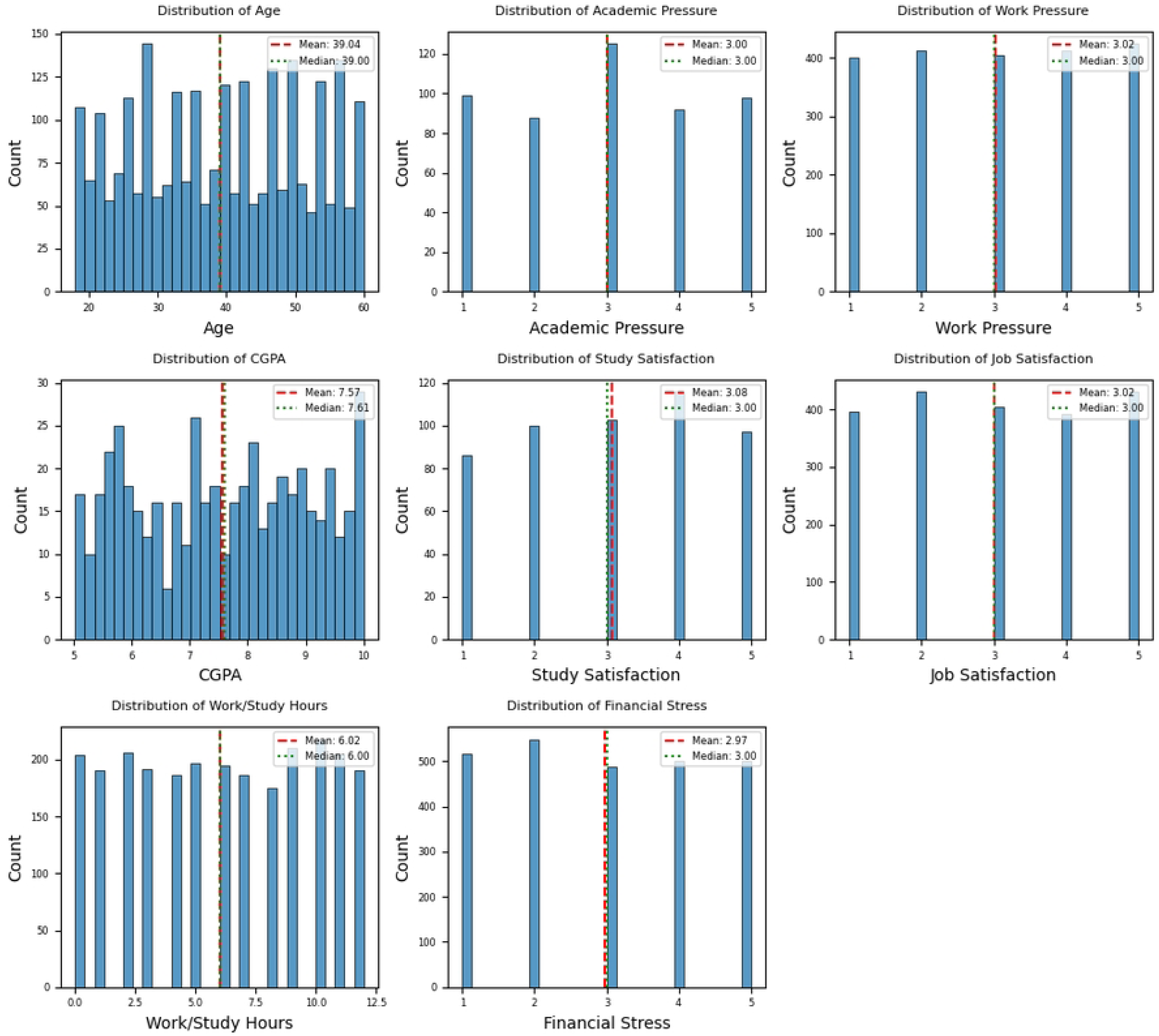

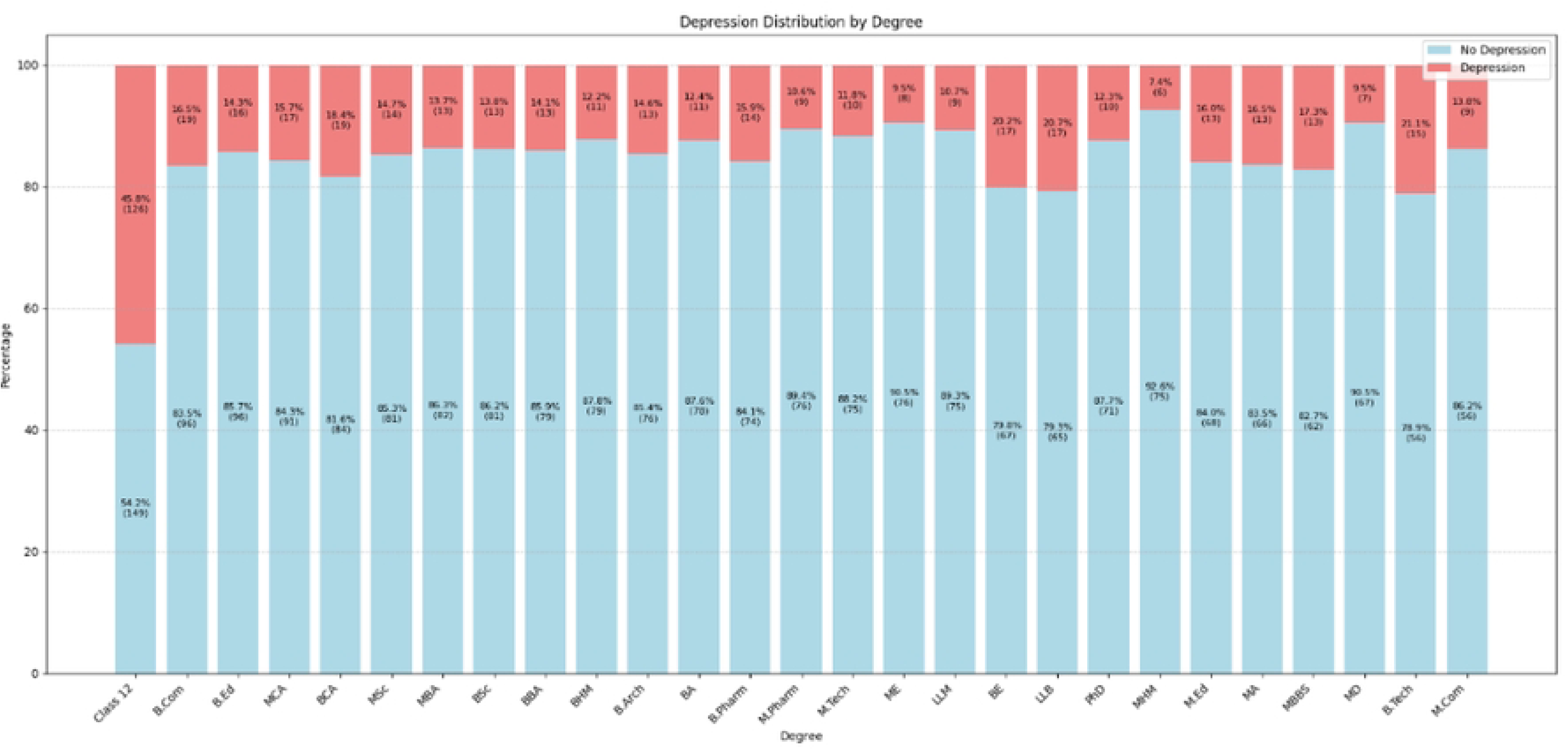

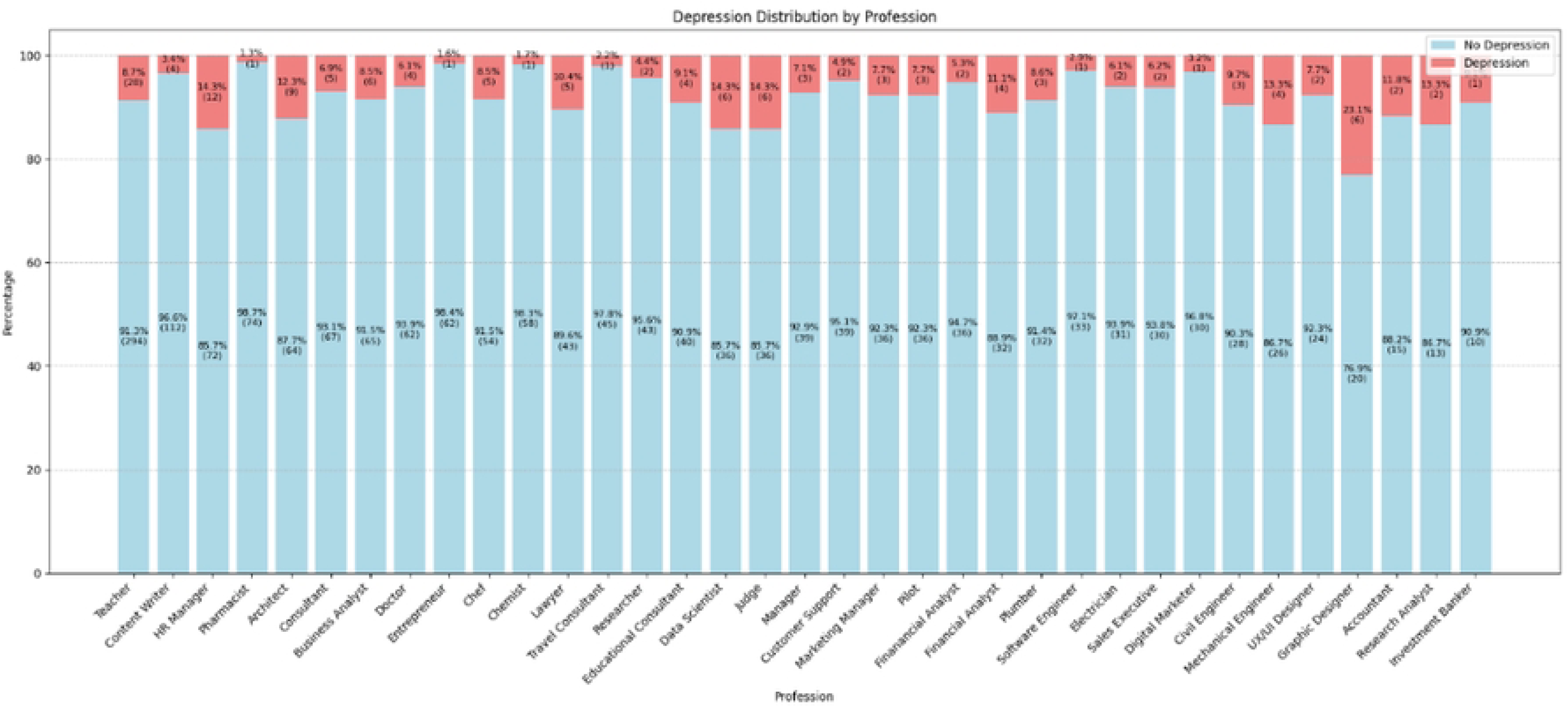

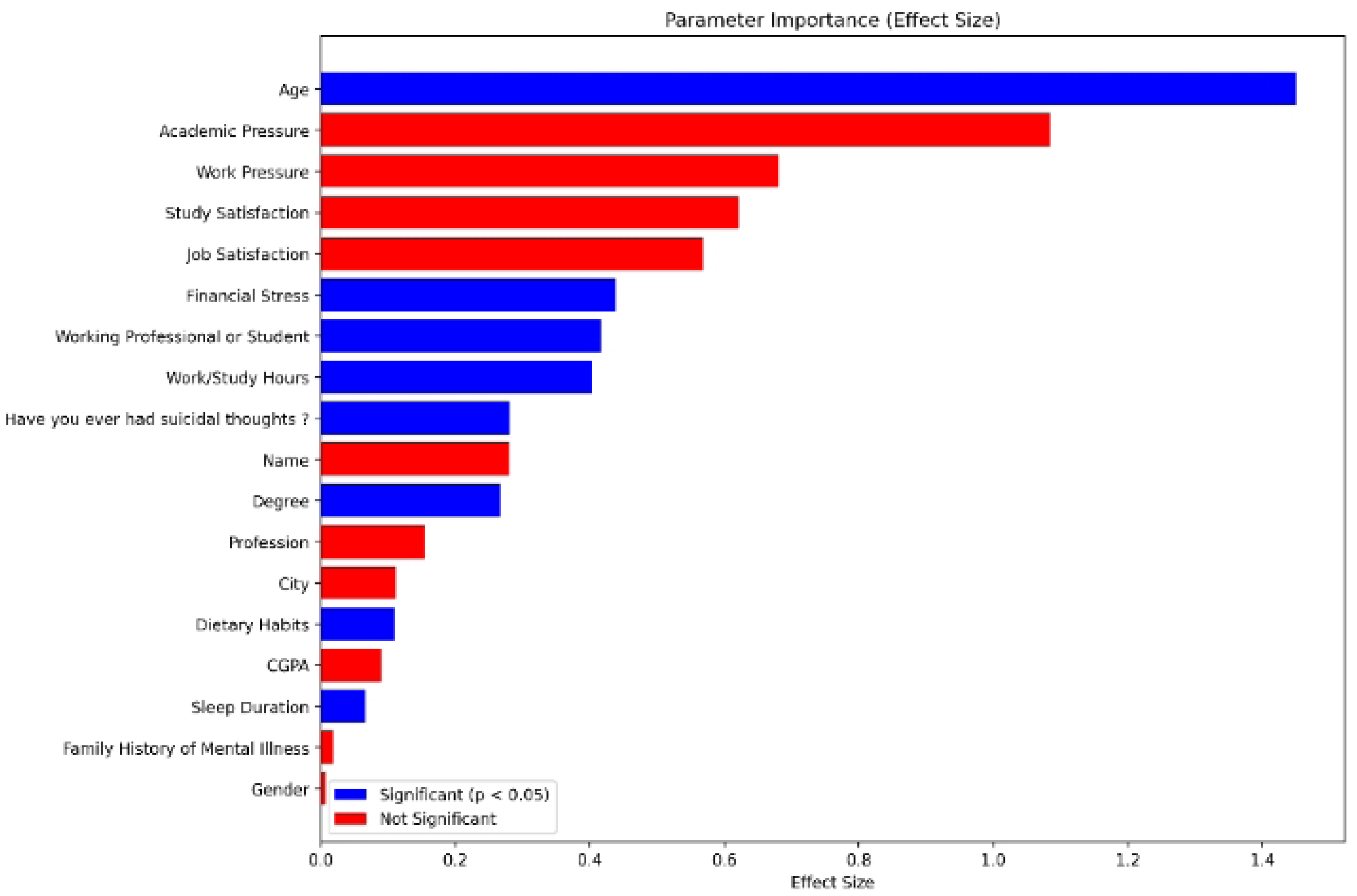

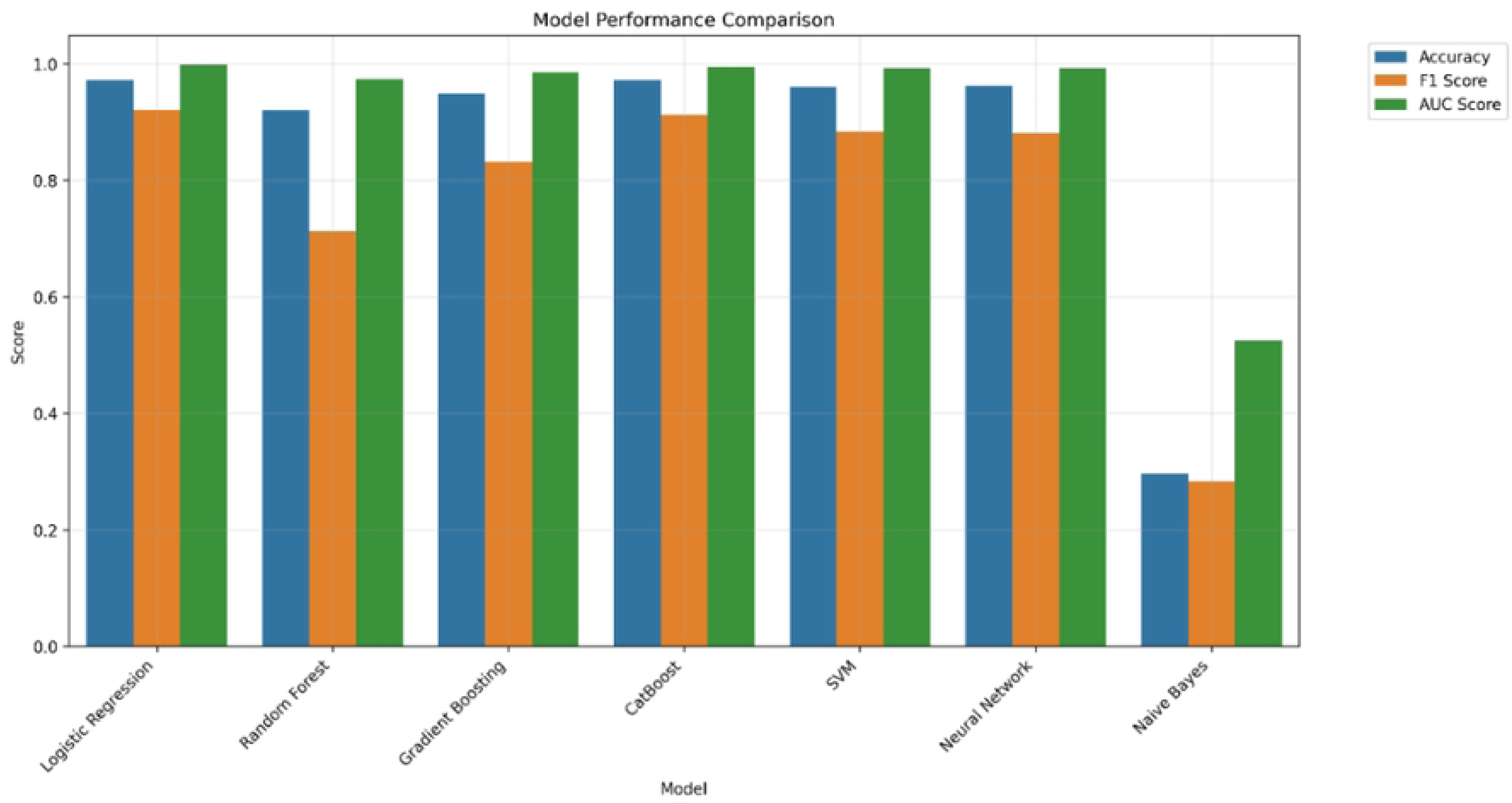

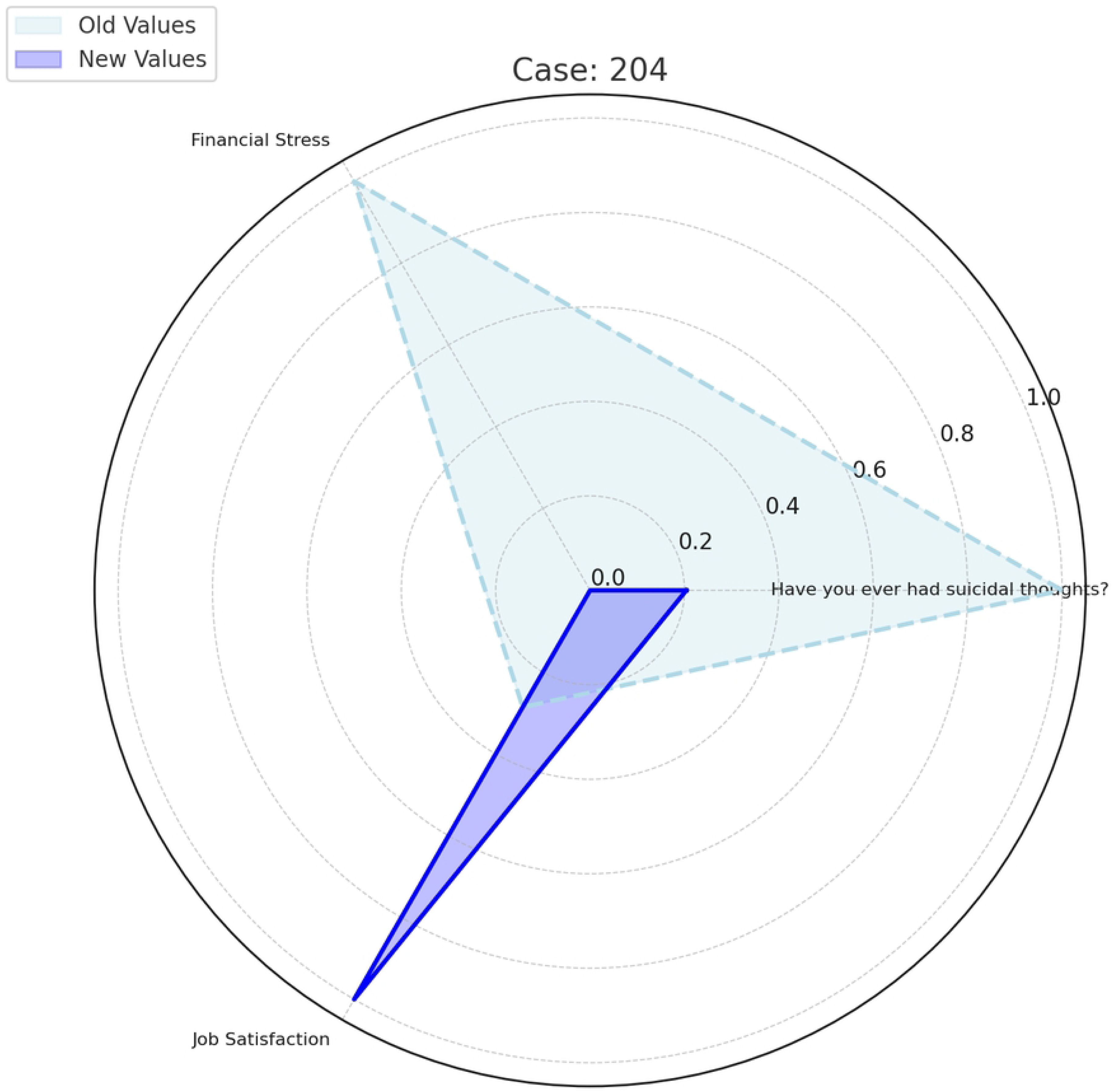

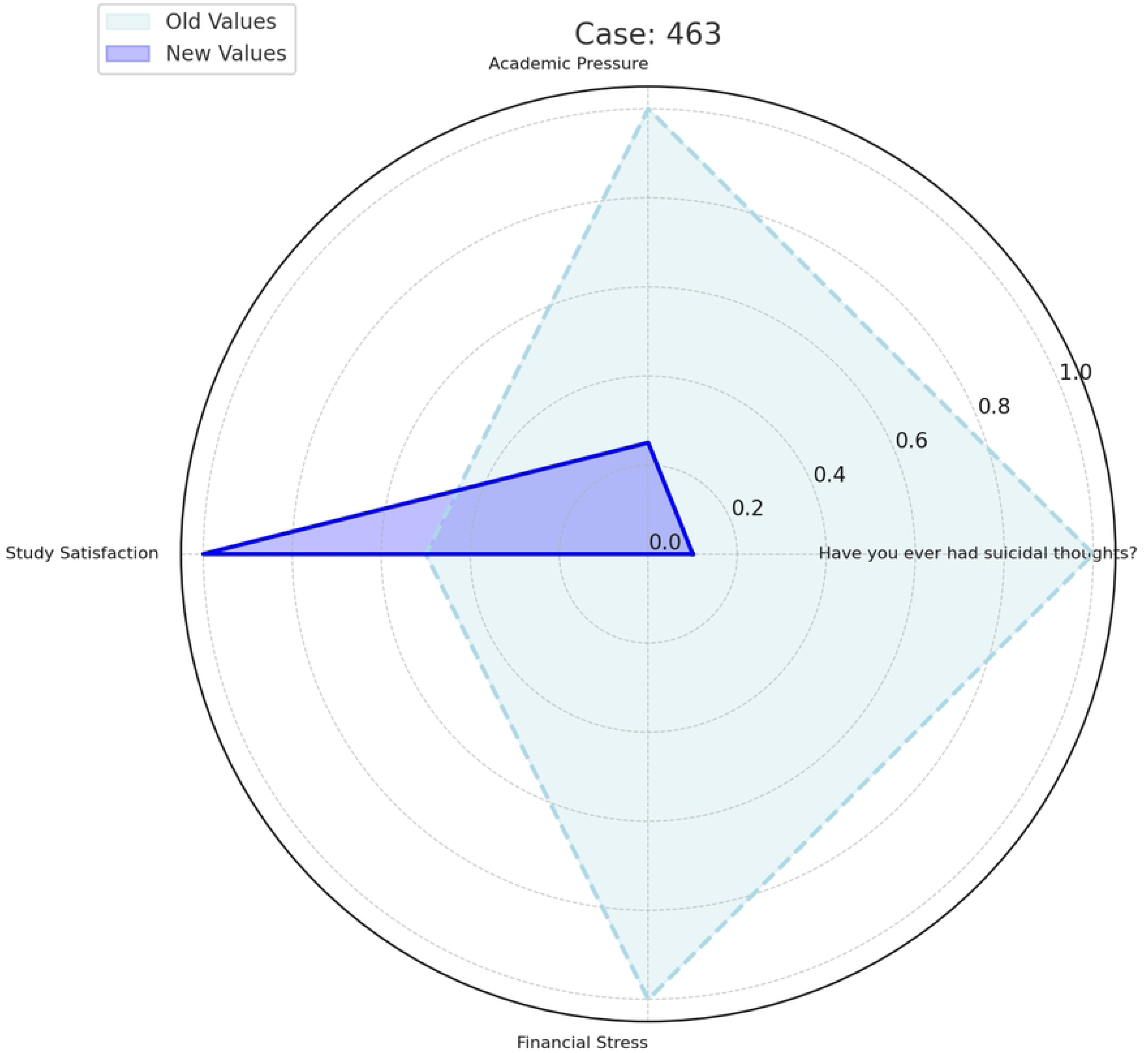

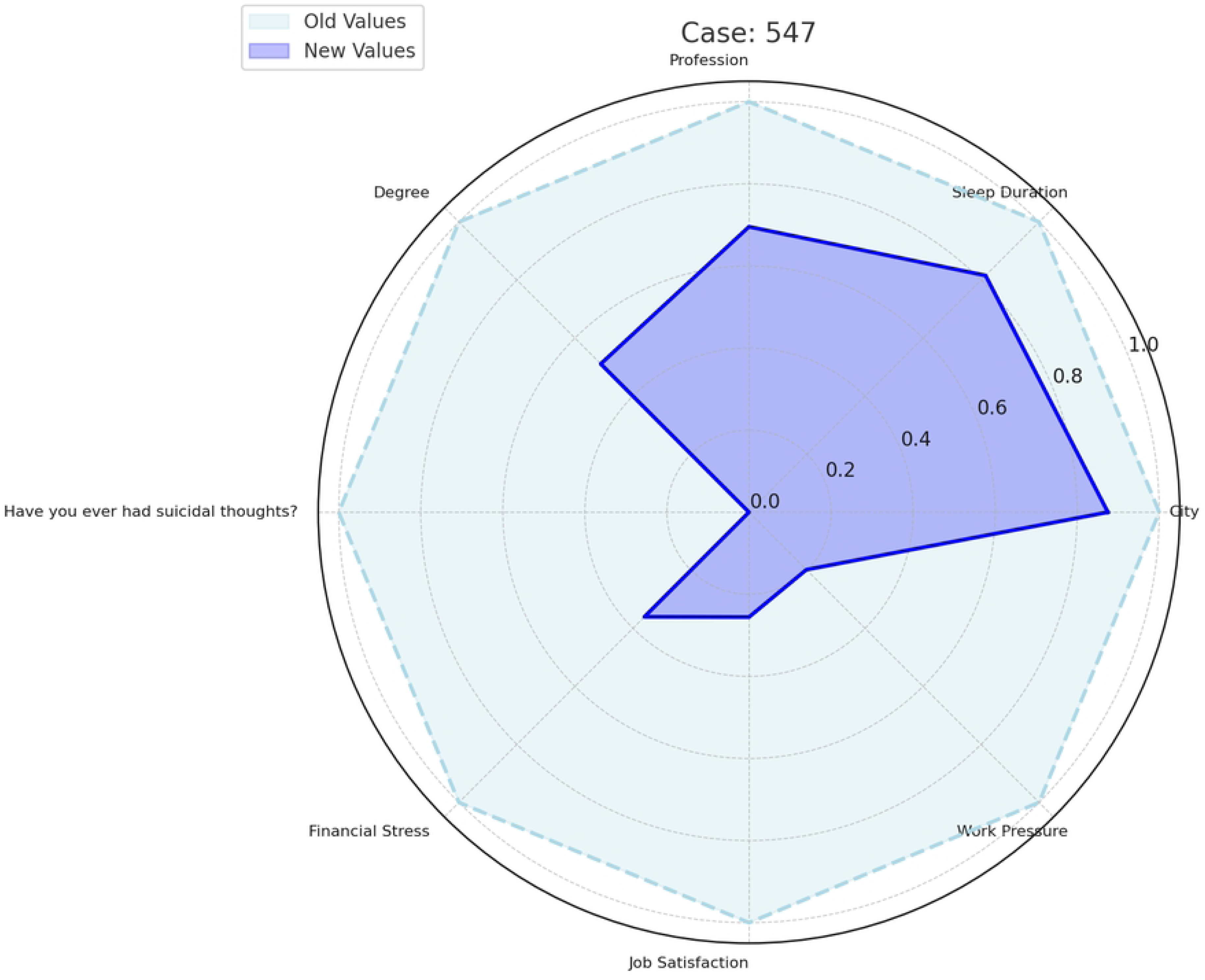

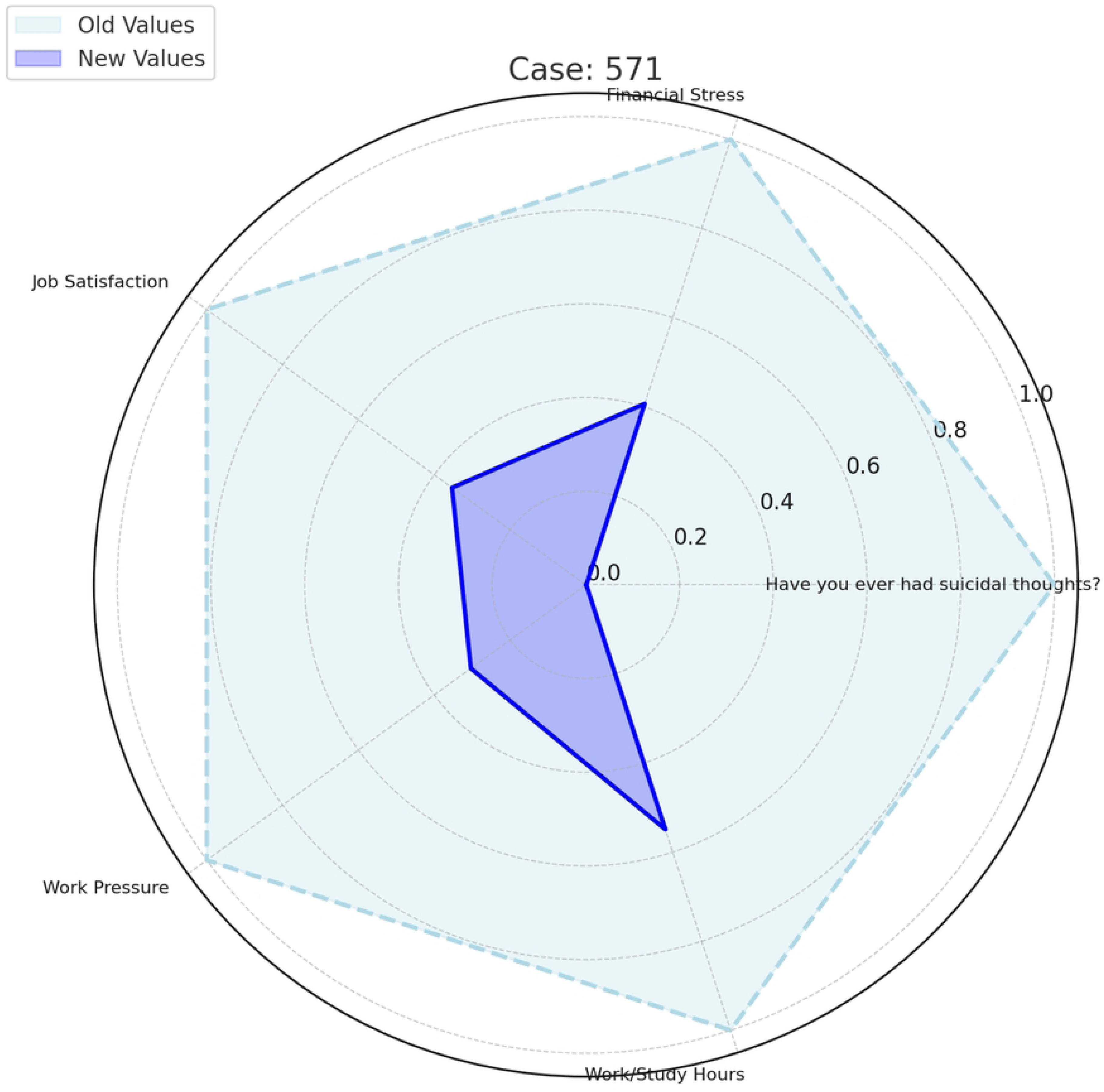

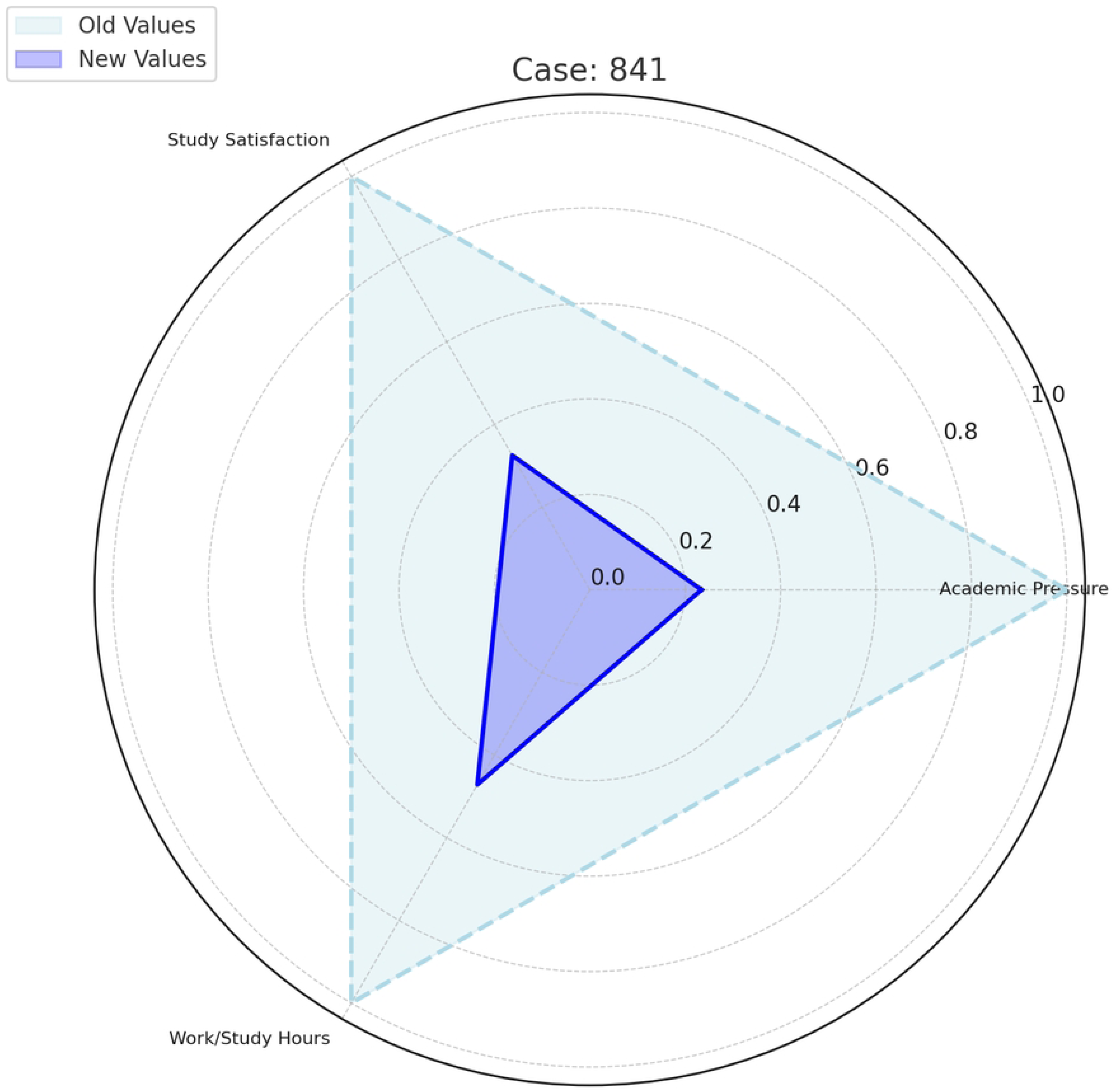

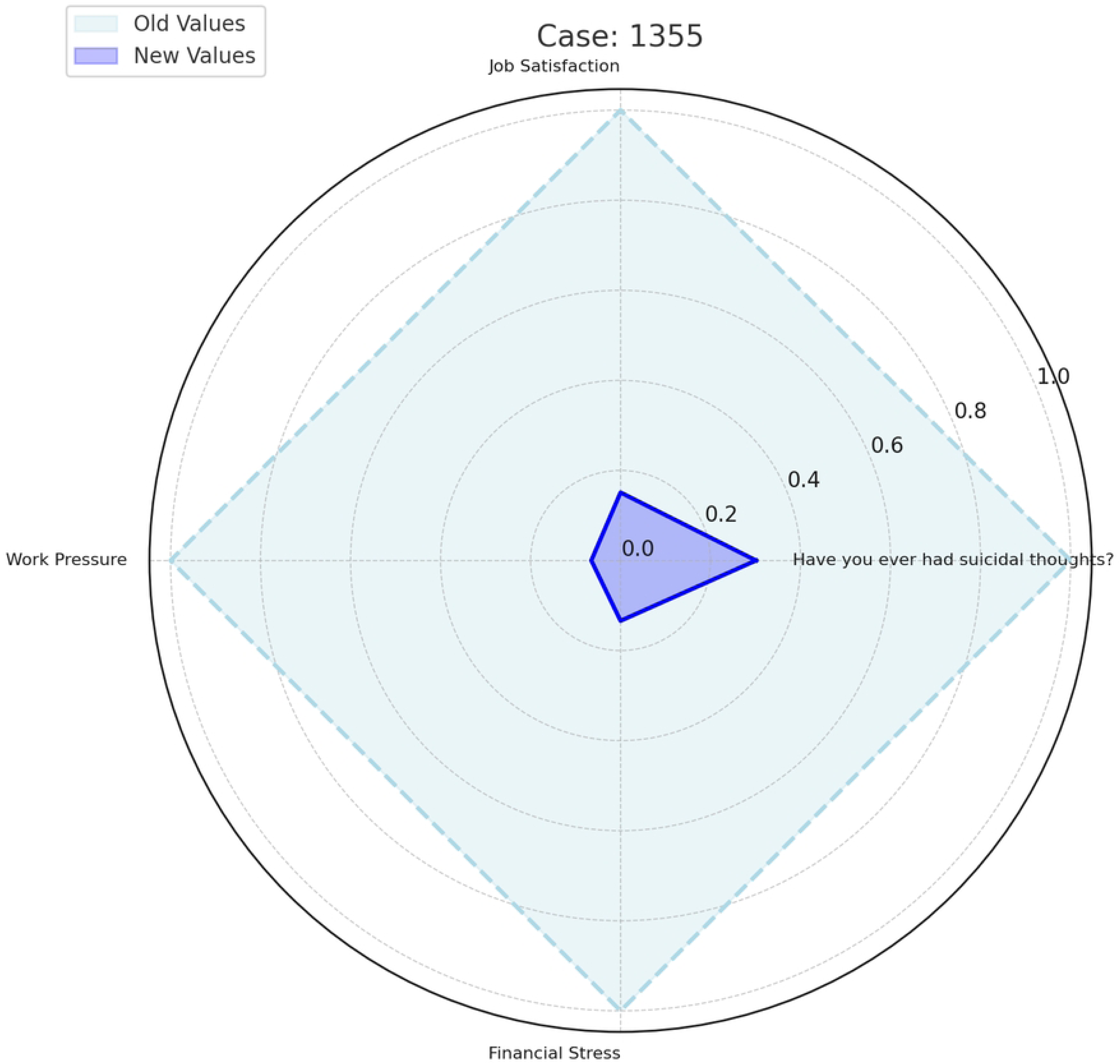

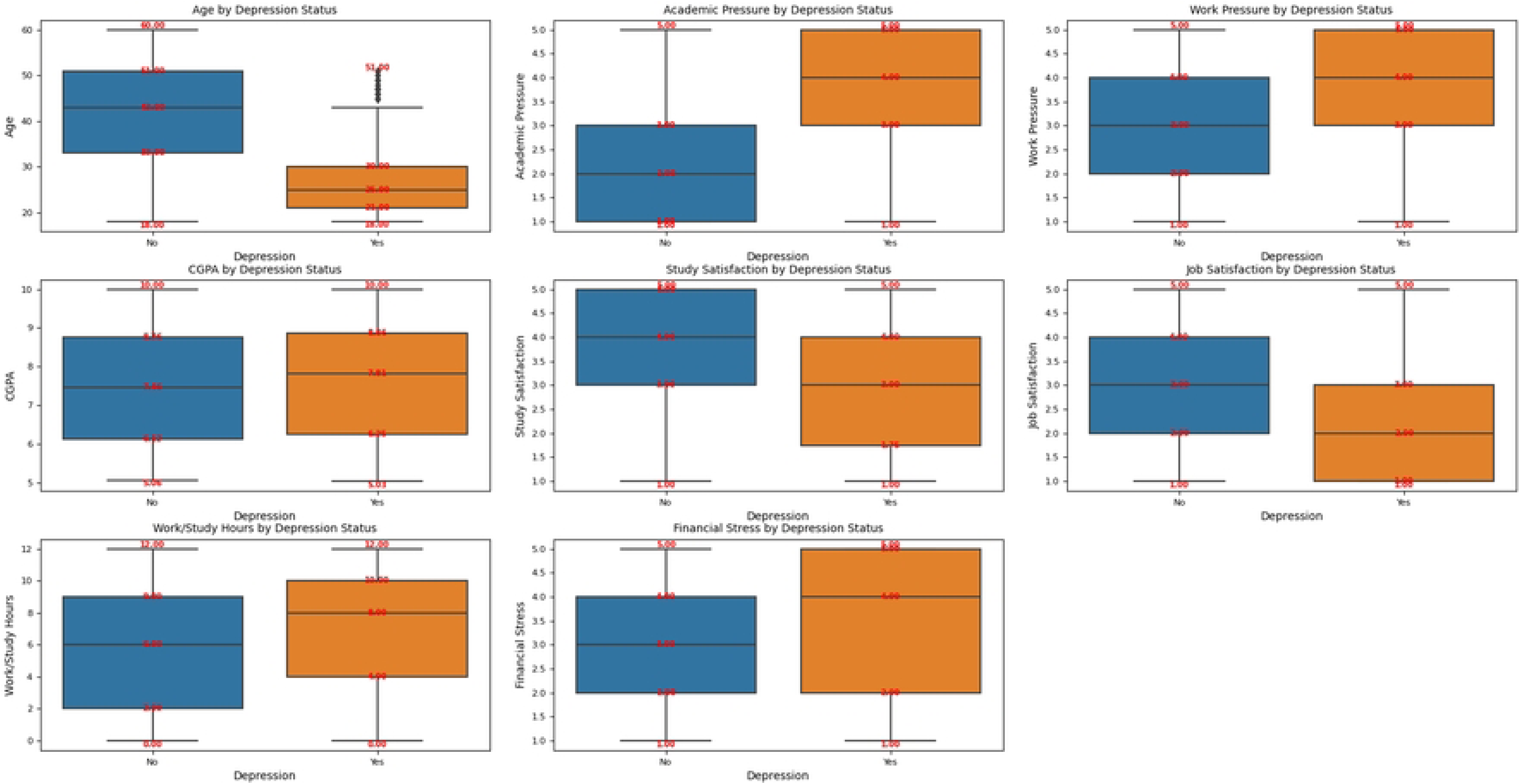

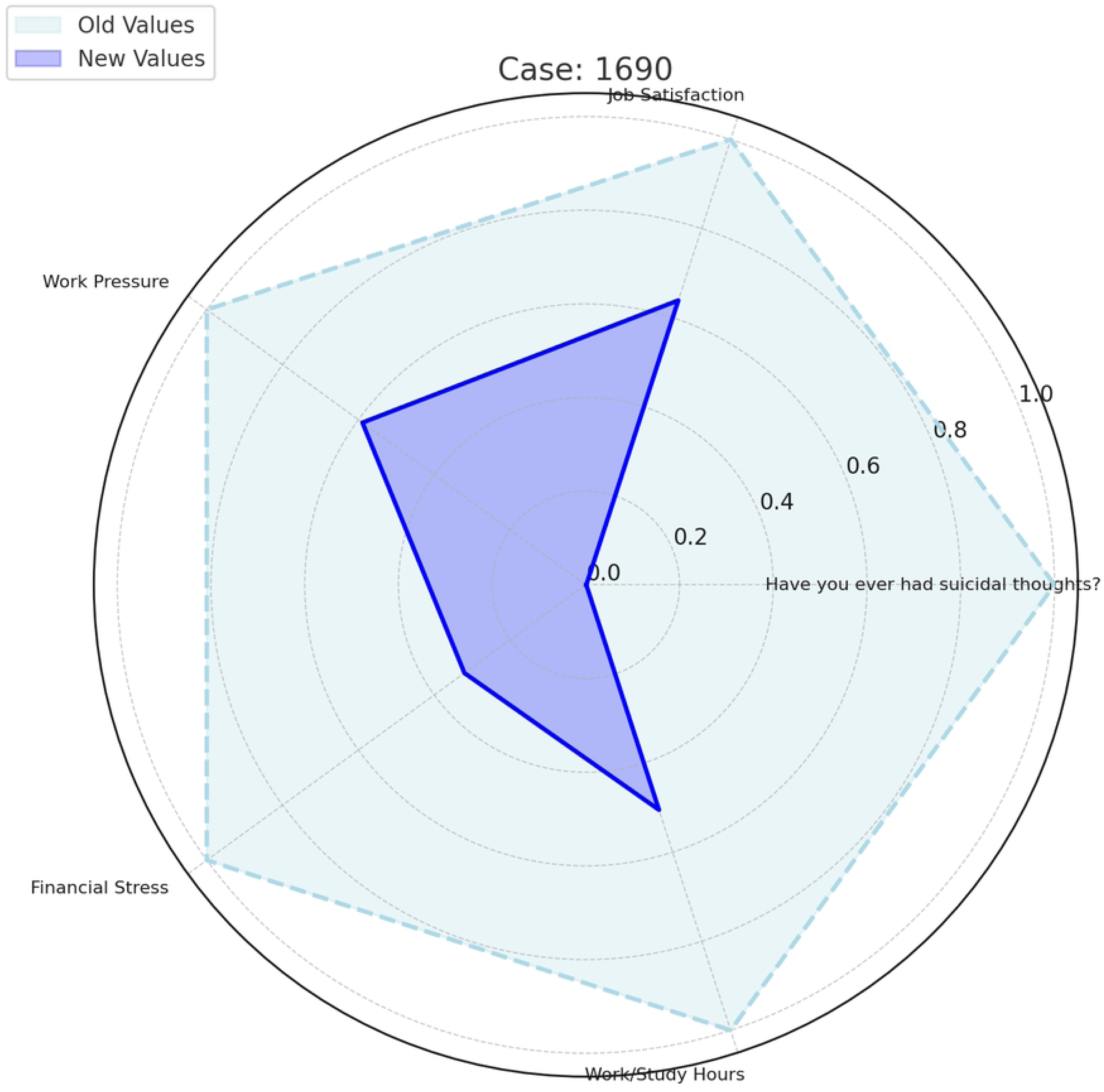

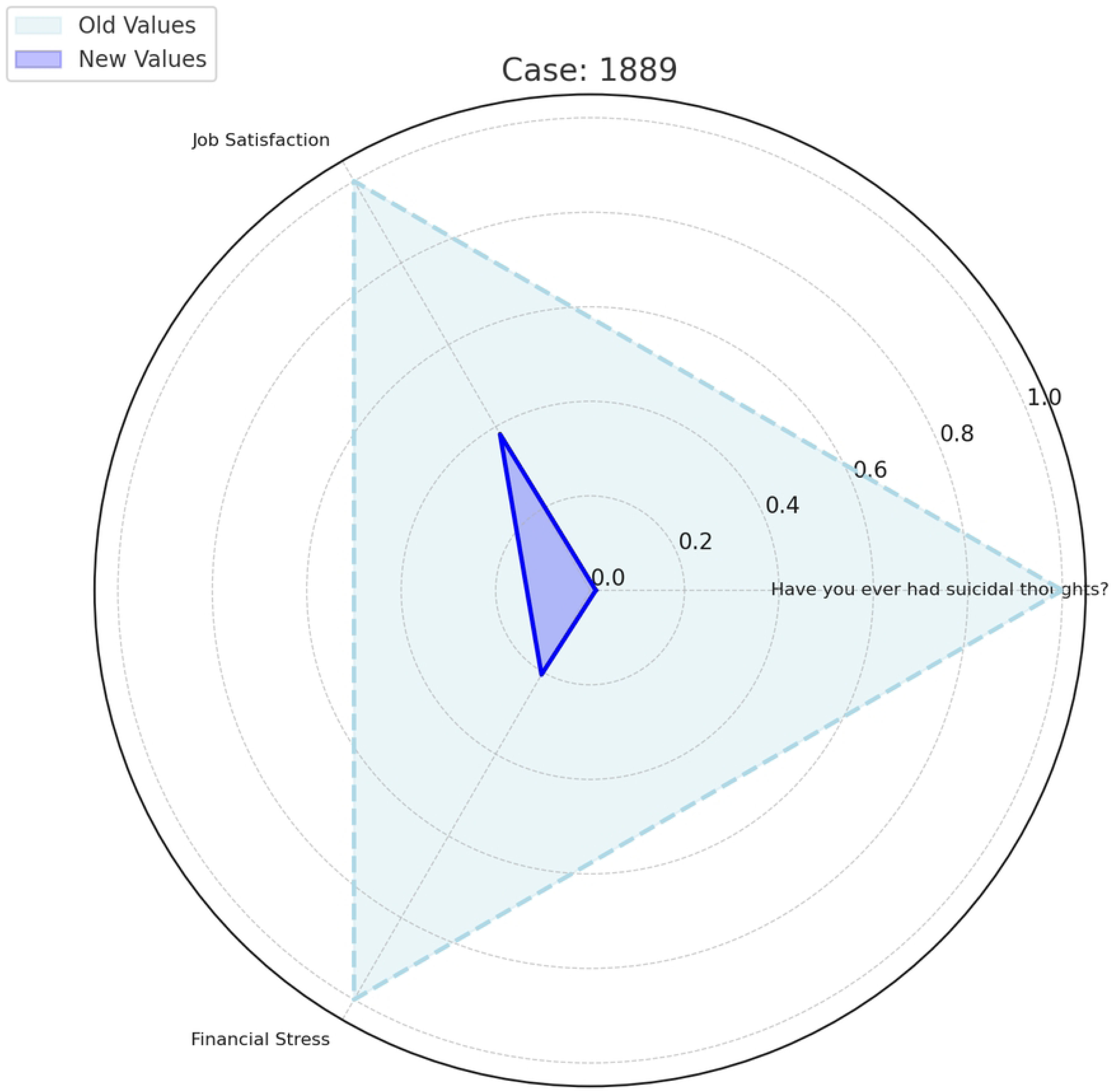

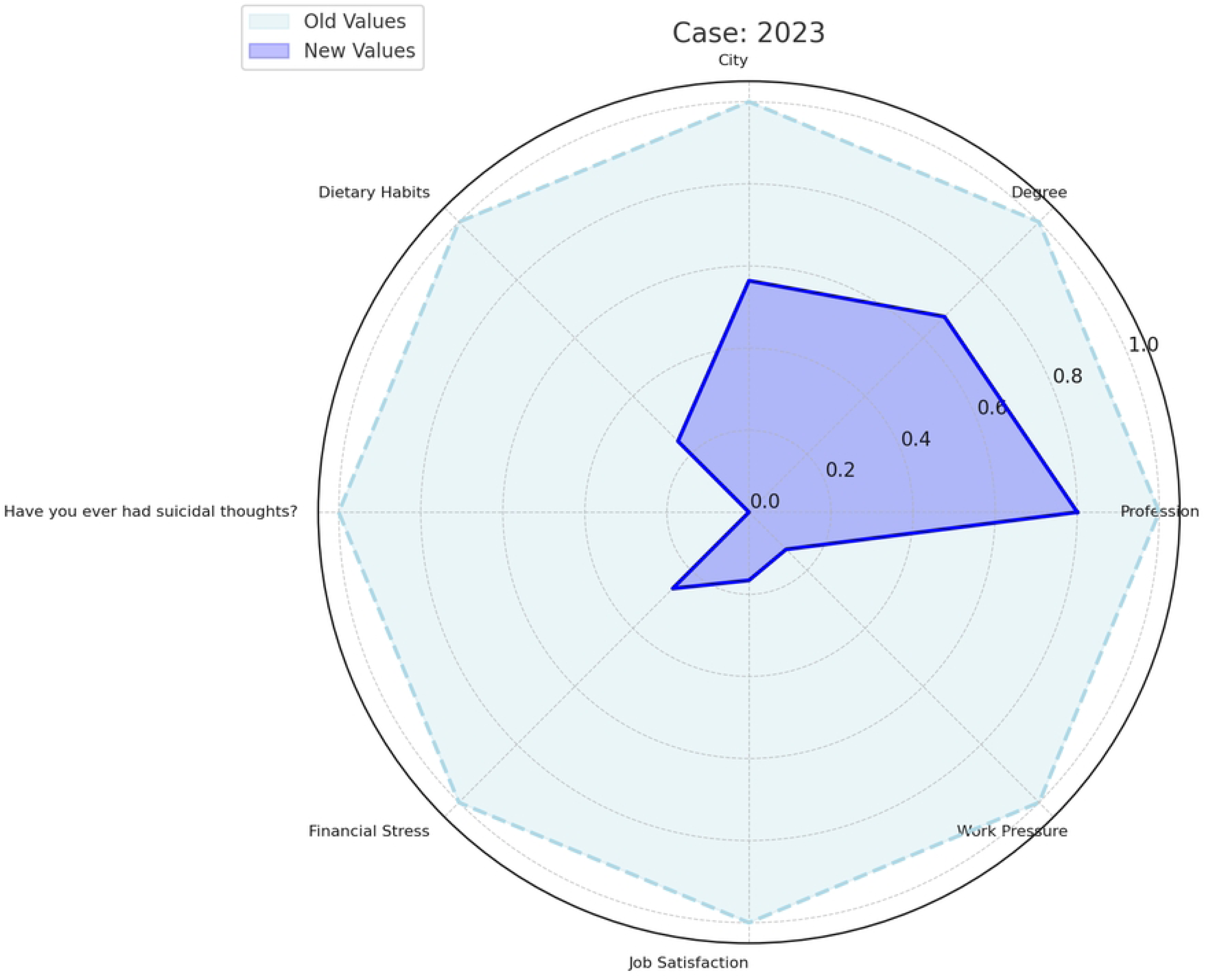

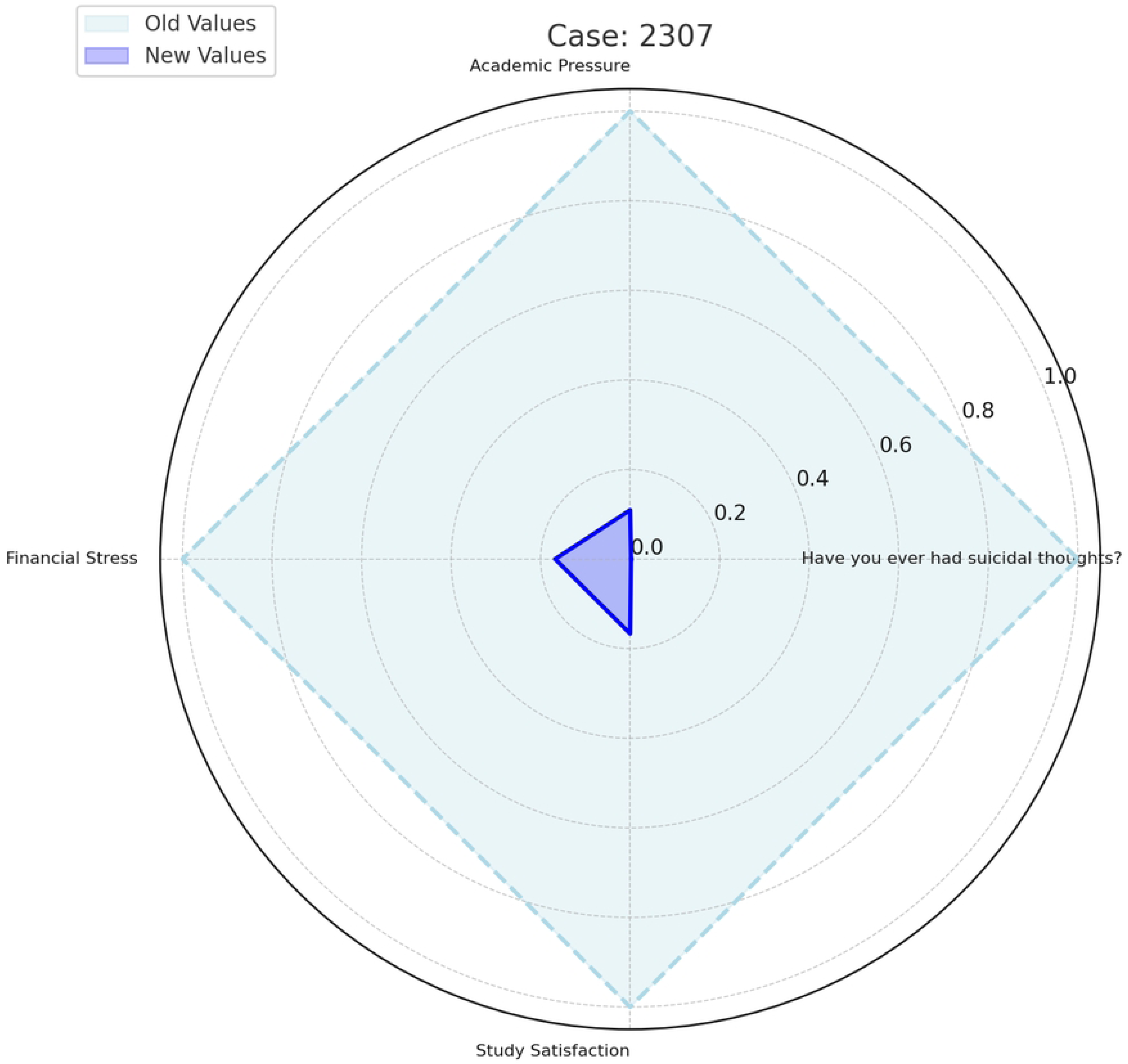

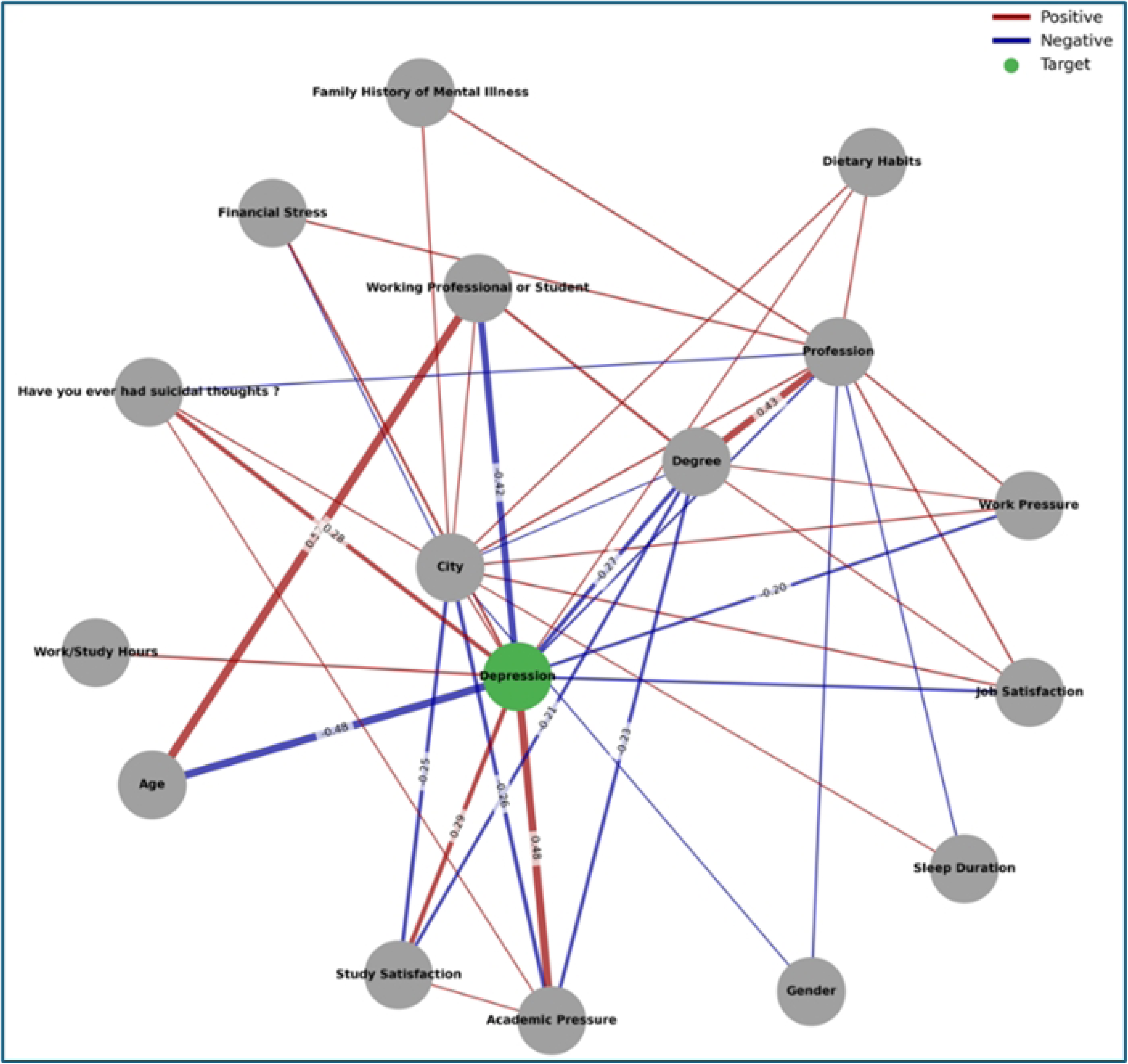

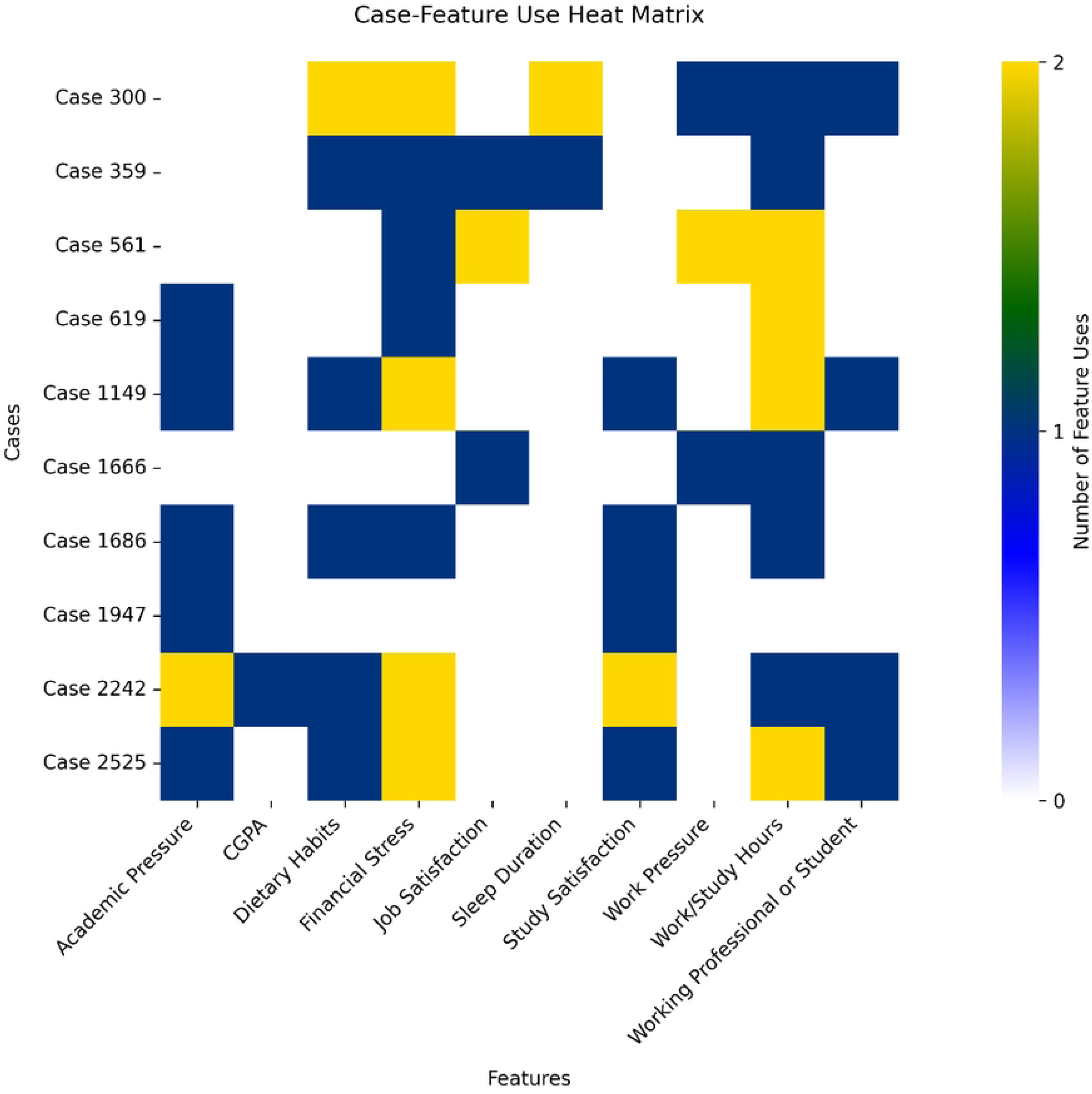

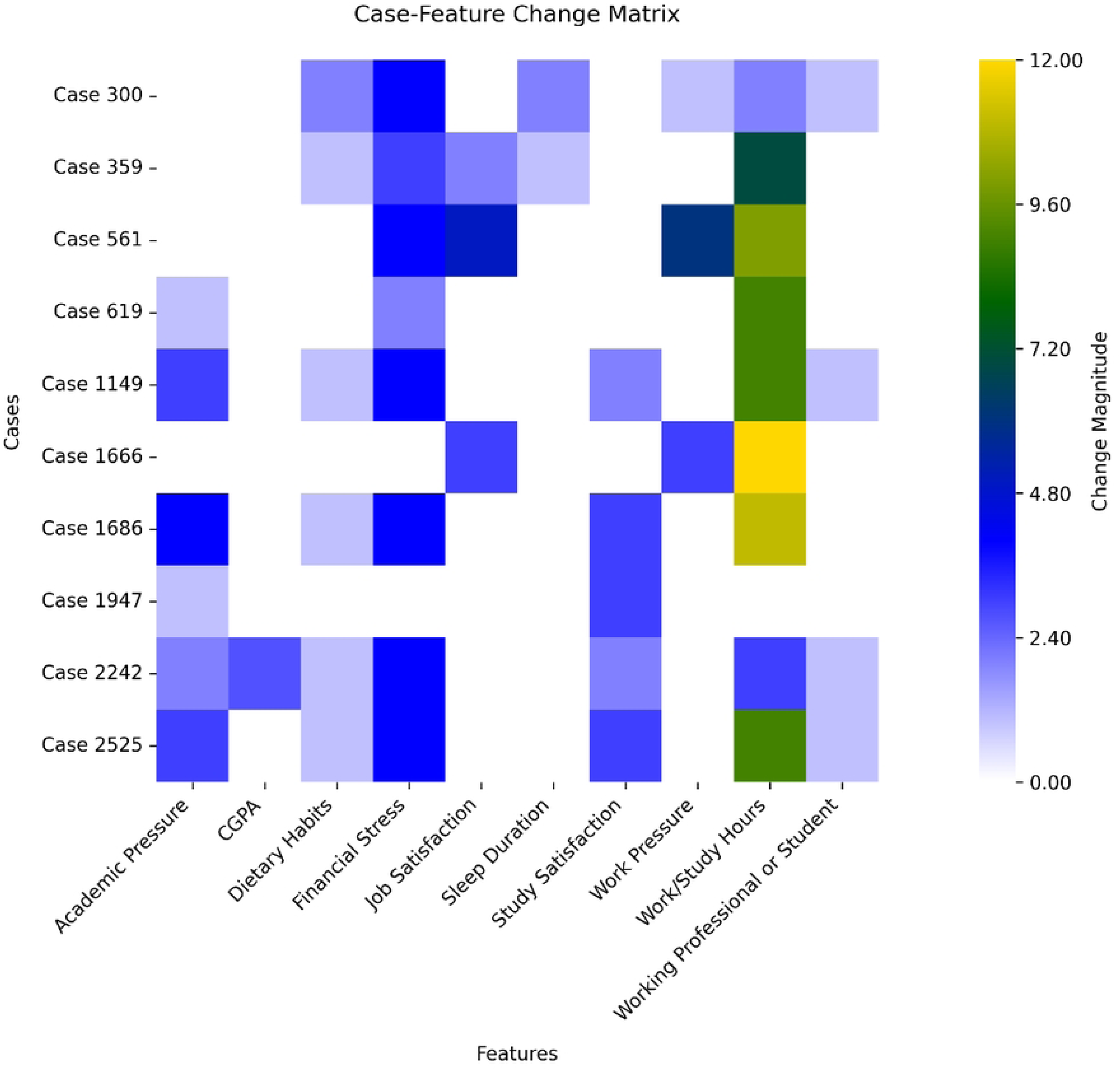

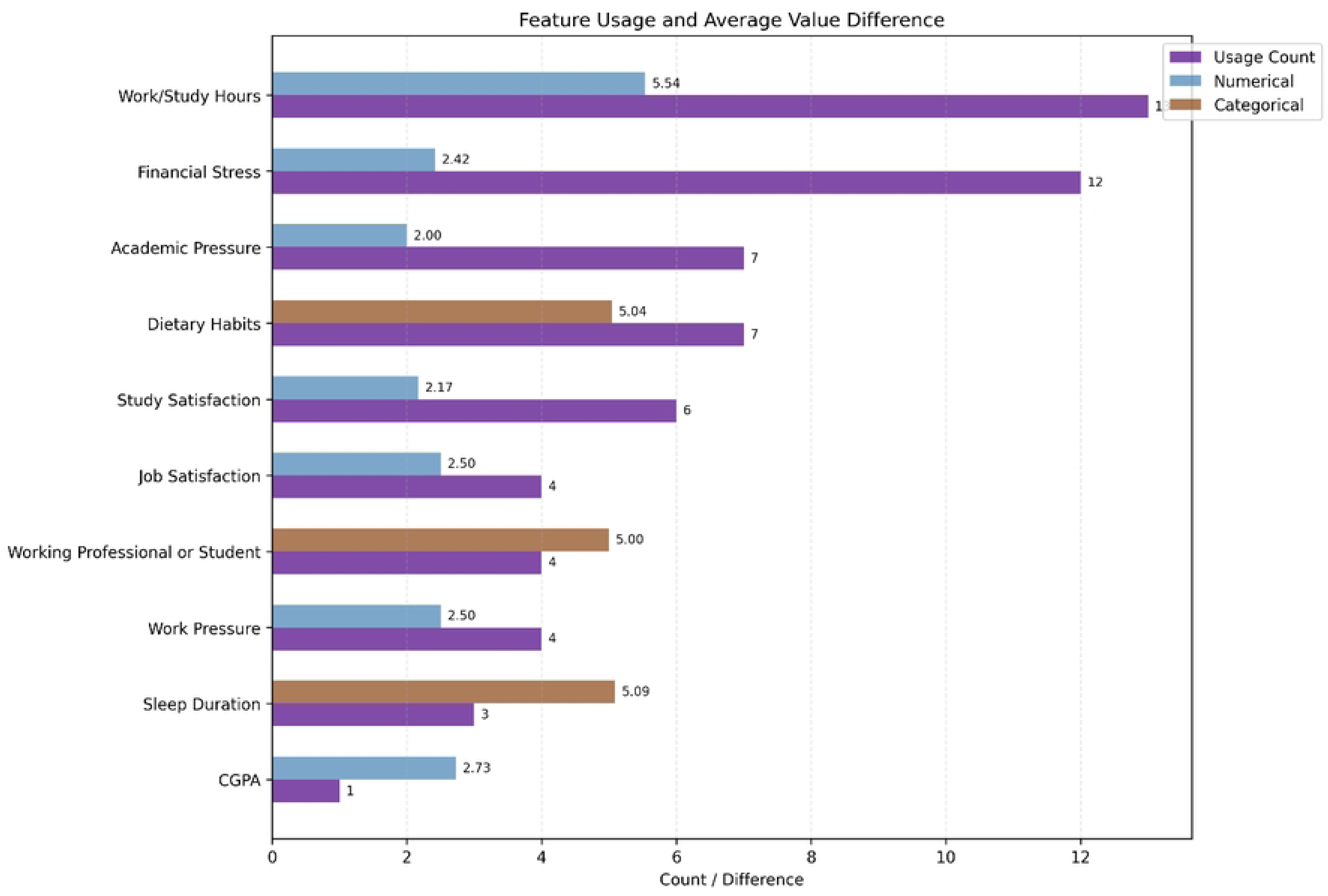

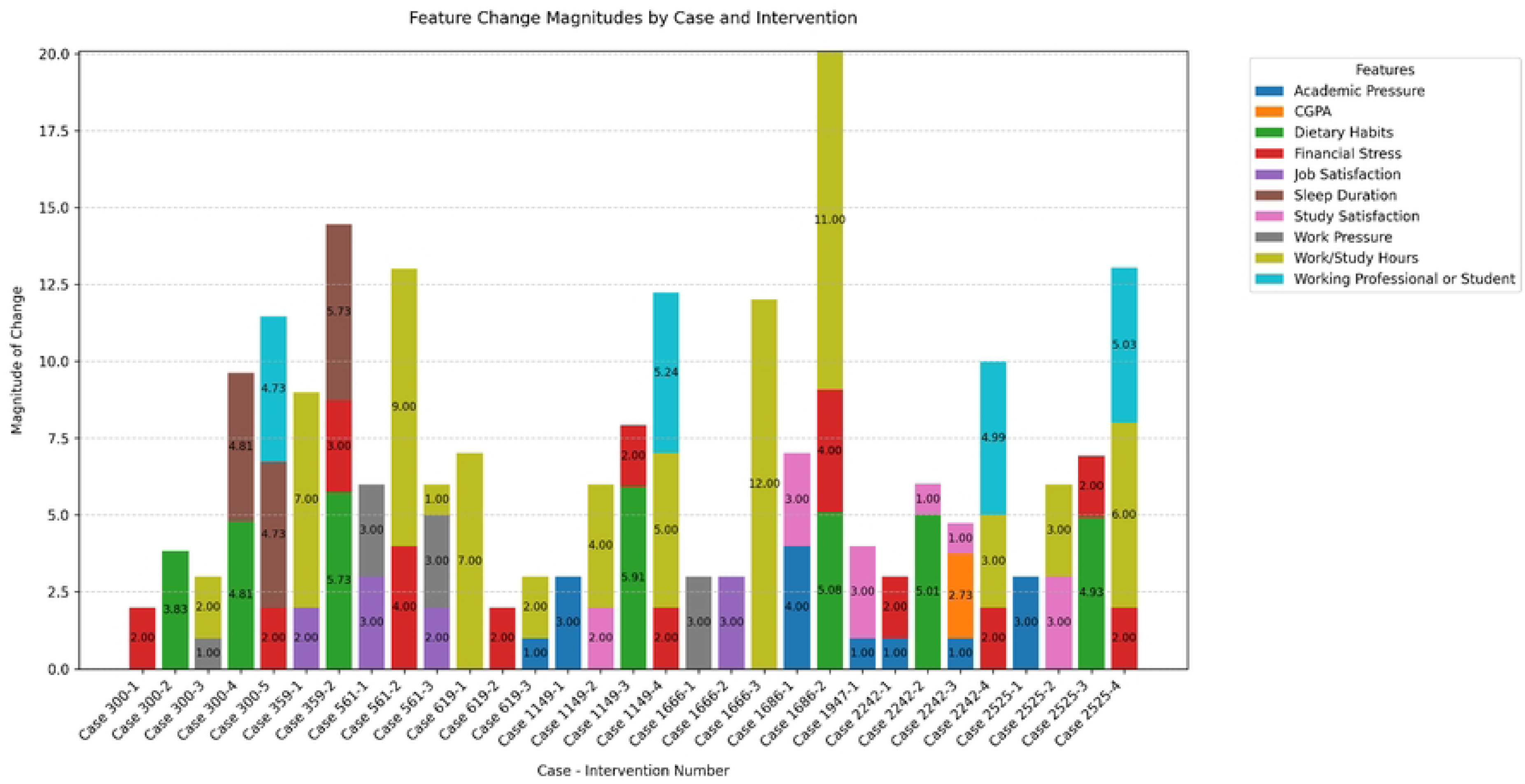

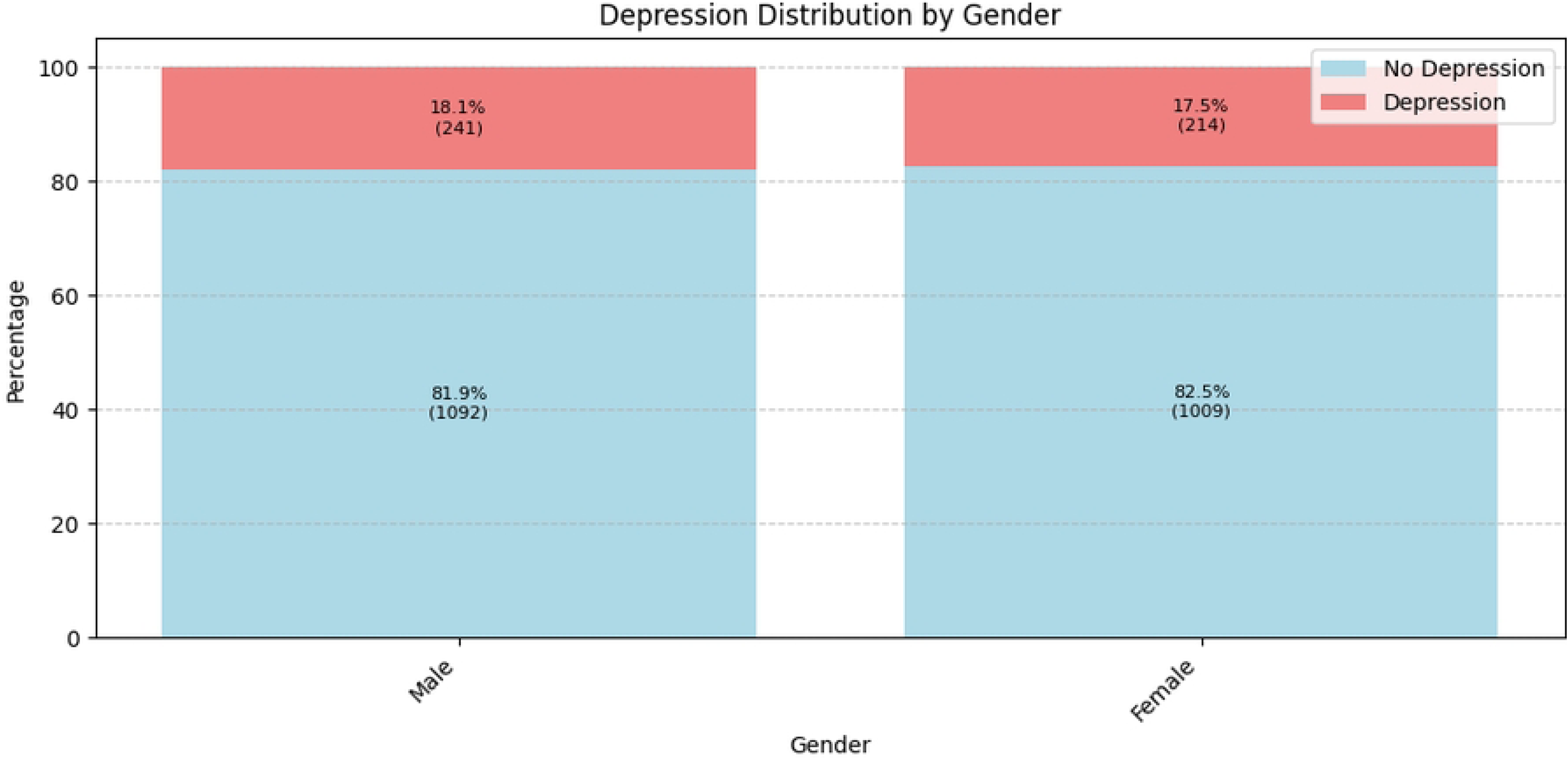

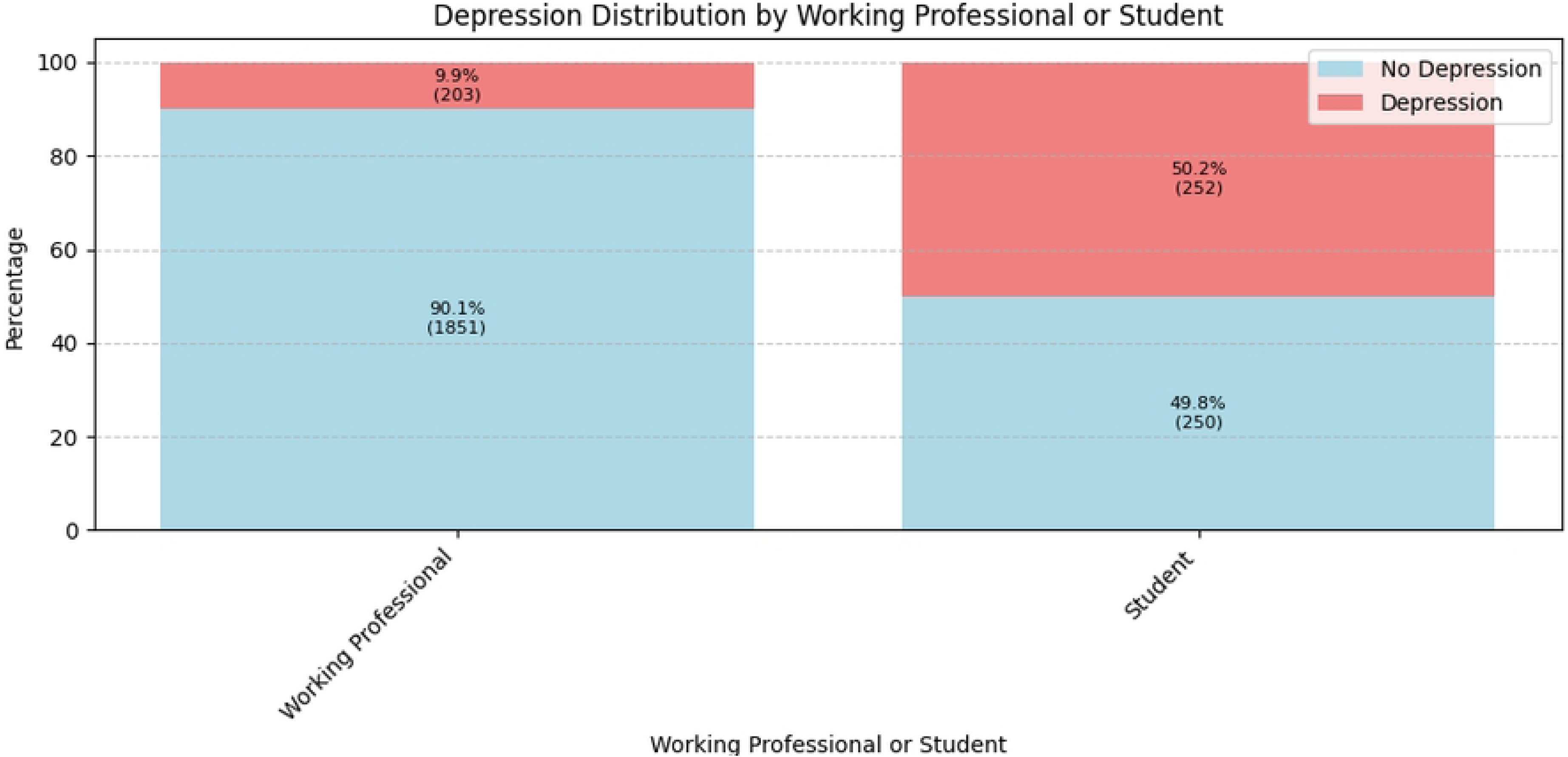

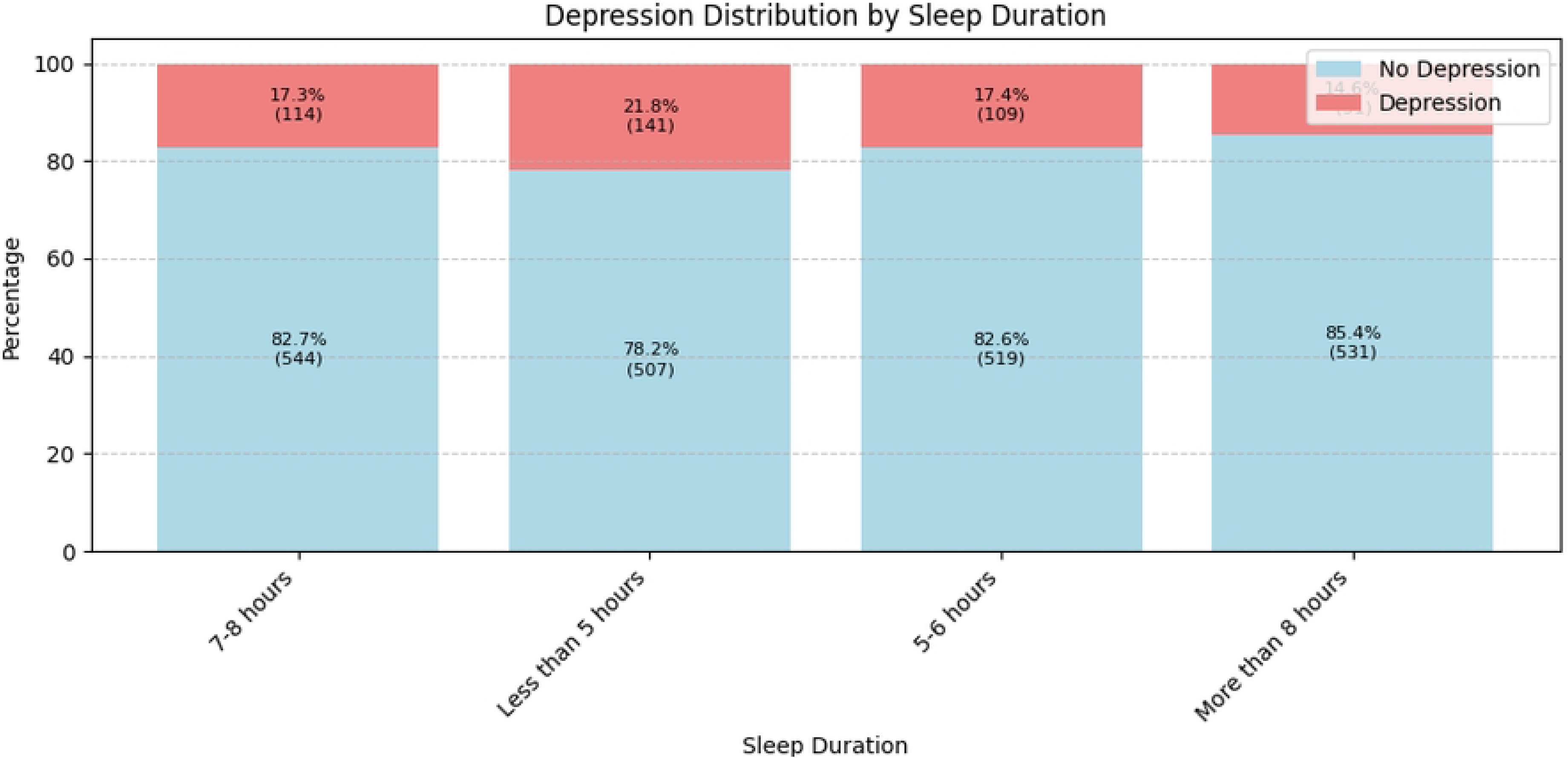

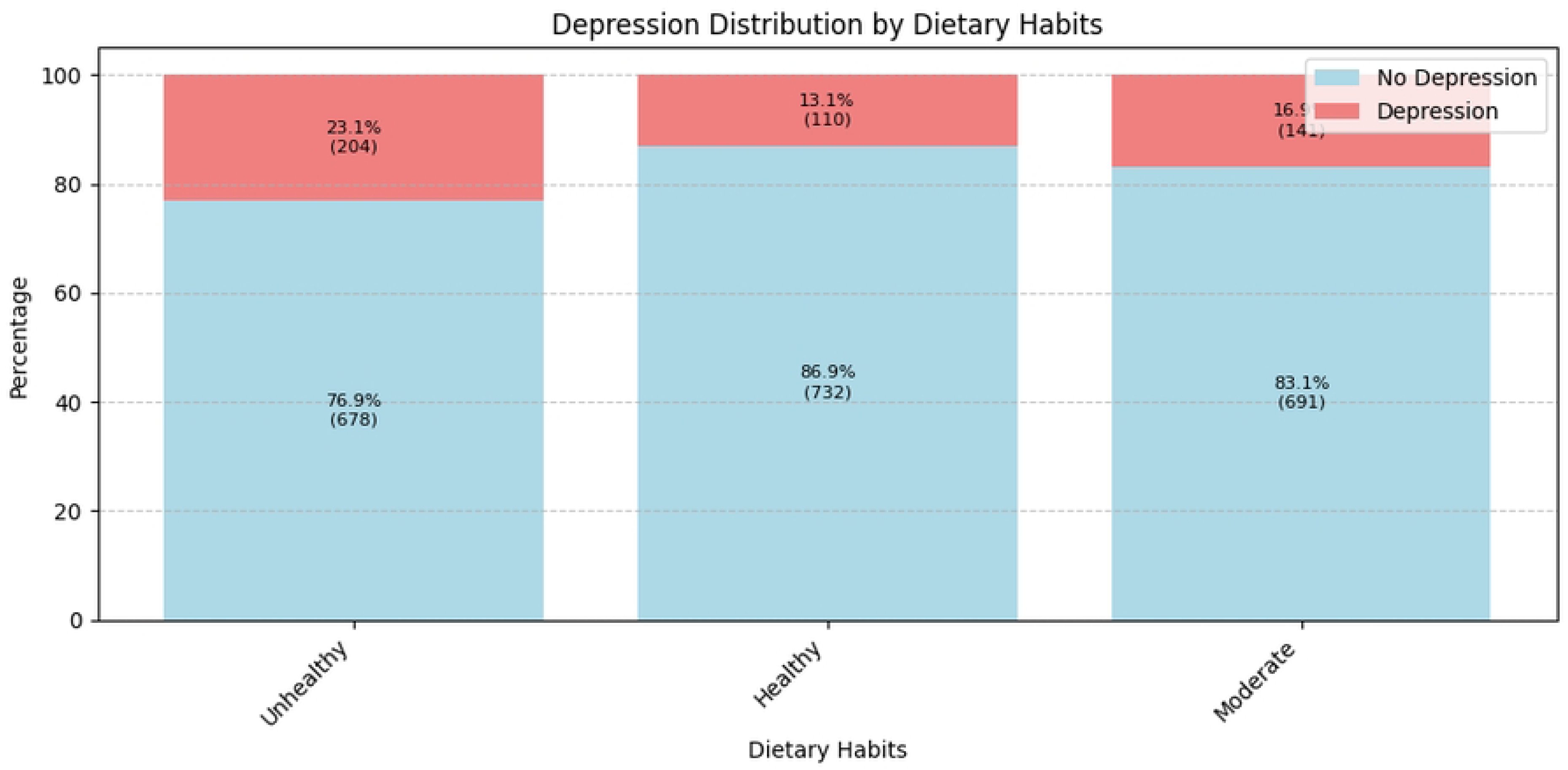

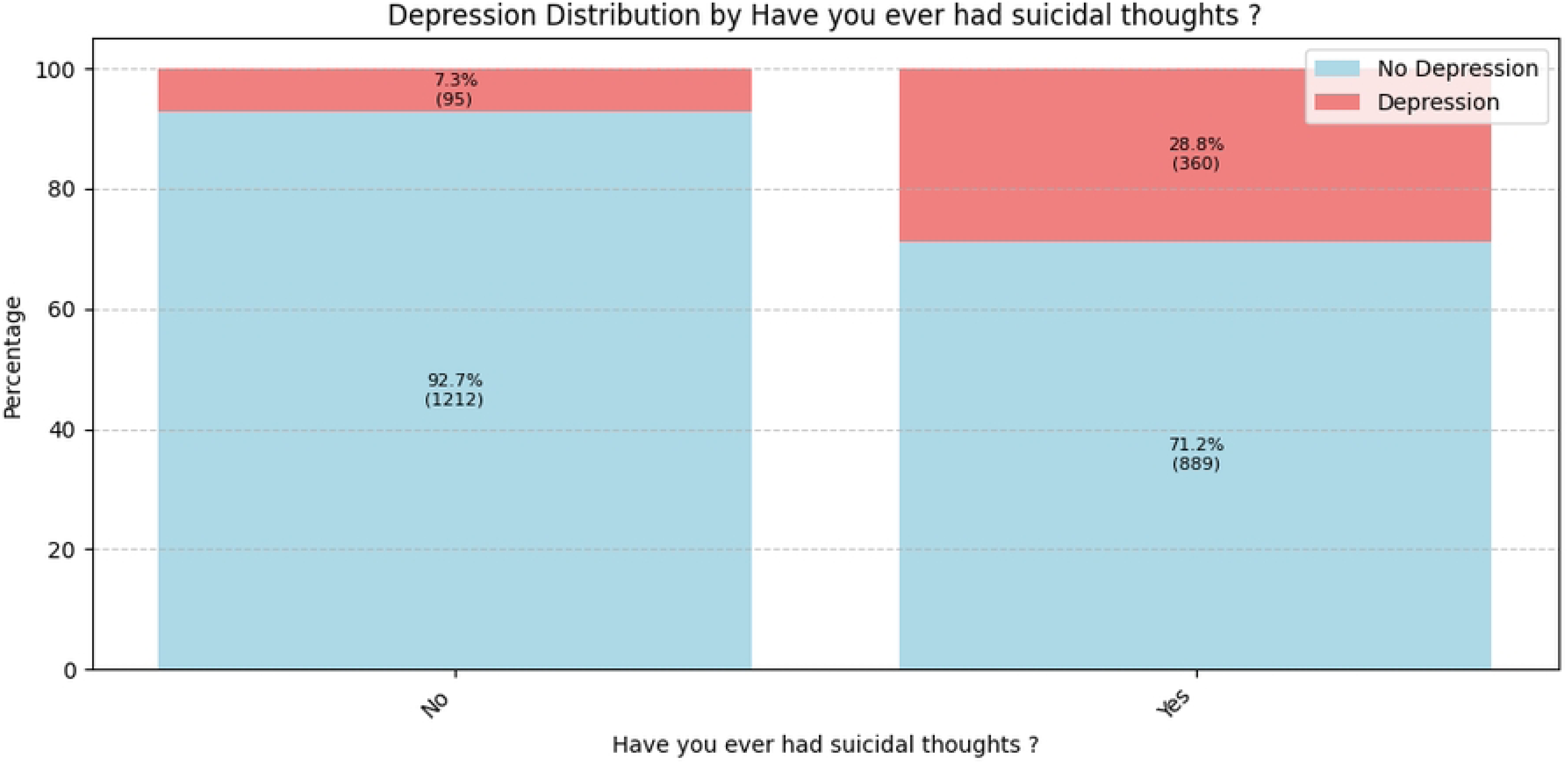

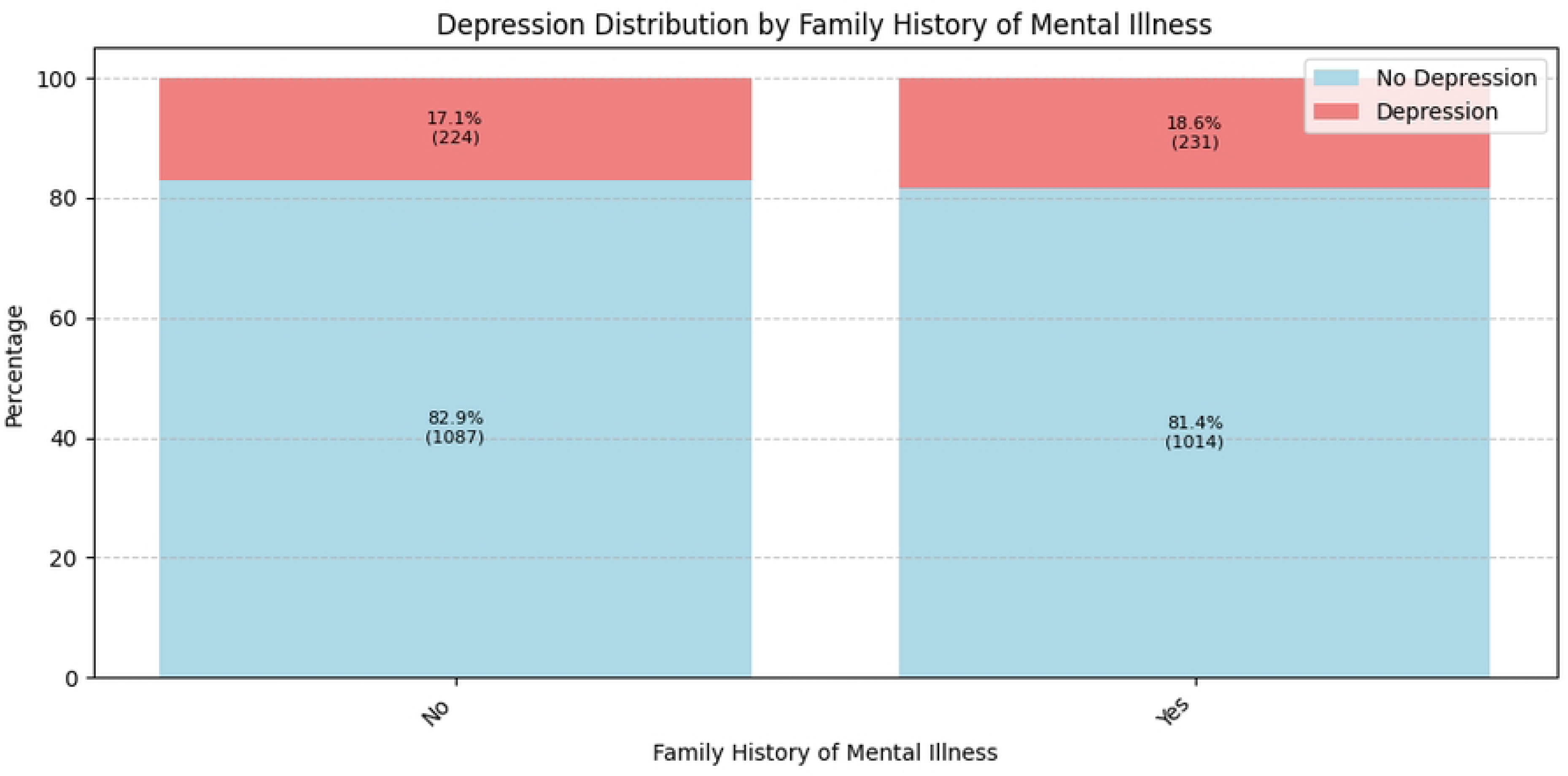

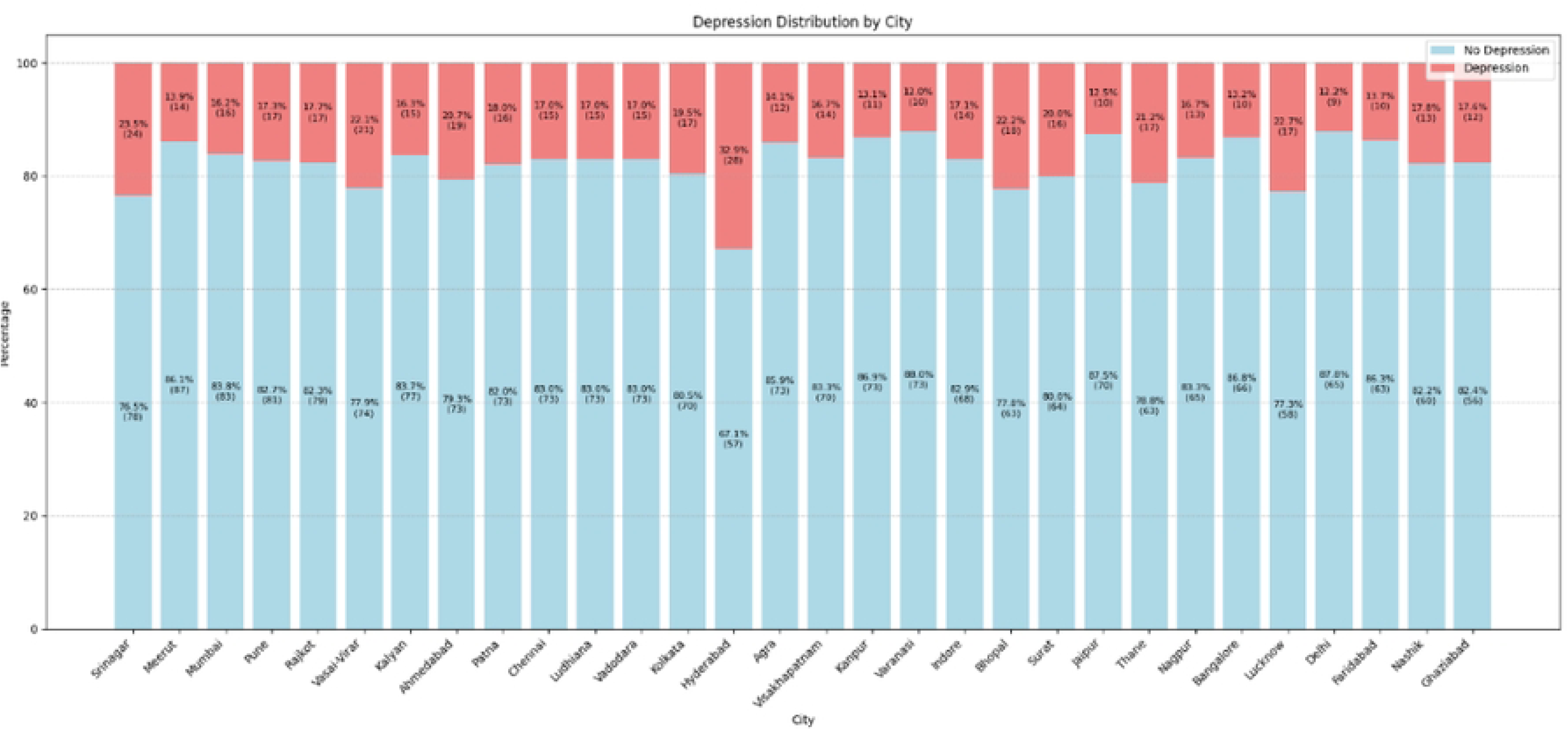

